# Genetic overlap between multivariate measures of human functional brain connectivity and psychiatric disorders

**DOI:** 10.1101/2021.06.15.21258954

**Authors:** Daniel Roelfs, Dennis van der Meer, Dag Alnæs, Oleksandr Frei, Alexey A. Shadrin, Robert Loughnan, Chun Chieh Fan, Anders M. Dale, Ole A. Andreassen, Lars T. Westlye, Tobias Kaufmann

**Affiliations:** NORMENT, Division of Mental Health and Addiction, Oslo University Hospital & Institute of Clinical Medicine, University of Oslo, Oslo, Norway; School of Mental Health and Neuroscience, Faculty of Health, Medicine, and Life Sciences, Maastricht University, Maastricht, the Netherlands; Center for Bioinformatics, Department of Informatics, University of Oslo, Oslo, Norway; Department of Cognitive Science, University of California, San Diego, 9500 Gilman Drive, La Jolla, CA 92093 USA; Population Neuroscience and Genetics Lab, University of California, San Diego, CA, USA; Center for Human Development, University of California, San Diego, CA, USA; Department of Radiology, School of Medicine, University of California, San Diego, CA, USA; Department of Neurosciences, University of California San Diego, La Jolla, CA 92037, USA; Center for Multimodal Imaging and Genetics, University of California at San Diego, La Jolla, CA, 92037, USA; K.G. Jebsen Center for Neurodevelopmental disorders, University of Oslo, Oslo, Norway; Department of Psychology, University of Oslo, Oslo, Norway; Department of Psychiatry and Psychotherapy, Tübingen Center for Mental Health, University of Tübingen, Germany; German Center for Mental Health (DZPG), partner site Tübingen, Germany

## Abstract

Psychiatric disorders are complex, heritable, and highly polygenic. Supported by findings of abnormalities in functional magnetic resonance imaging (fMRI) based measures of brain connectivity, current theoretical and empirical accounts have conceptualized them as disorders of brain connectivity and dysfunctional integration of brain signaling, however, the extent to which these findings reflect common genetic factors remains unclear. Here, we performed a multivariate genome-wide association analysis of fMRI-based functional brain connectivity in a sample of 30,701 individuals from the UK Biobank and investigated the shared genetic determinants with eight major psychiatric disorders. The analysis revealed significant genetic overlap between functional brain connectivity and schizophrenia, bipolar disorder, attention-deficit hyperactivity disorder, autism spectrum disorder, anxiety, and major depression, adding further genetic support for the dysconnectivity hypothesis of psychiatric disorders and identifying potential genetic and functional targets for future studies.

## Introduction

Psychiatric disorders are heritable and highly polygenic^1–4^, and carry a high burden of disease, measured in years lived with disability^5^. Akin to the polygenic architecture of the disorders, where a number of variants each contribute with small effects, findings from imaging genetics studies have documented a distributed pattern of small effects across the genome for brain phenotypes derived from magnetic resonance imaging (MRI)^6^. Likewise, brain imaging studies of psychiatric disorders have revealed distributed anatomical and functional alterations across the brain, with a large body of literature indicating alterations in functional brain connectivity in individuals with a range of psychiatric disorders, including schizophrenia (SCZ; e.g. Petterson-Yeo et al., 2011^7^), bipolar disorder (BIP; e.g. Syan et al., 2018^8^), autism spectrum disorders (ASD; e.g. Hong et al.,2019^9^), attention-deficit hyperactivity disorder (ADHD; e.g. Gao et al., 2019^10^), major depression (MDD; e.g. Brakowski et al., 2017^11^), post-traumatic stress disorder (PTSD; e.g. Akiki et al., 2017^12^), anxiety disorders (ANX; e.g. Xu et al., 2019^13^), and anorexia nervosa (AN, e.g. Fuglset et al., 2016^14^).

Altered brain connectivity in psychiatric disorders might reflect changes in synaptic functioning. Evidence from induced pluripotent stem cell research shows that mutations relevant to psychiatric disorders cause synapse deficits^15^, genome-wide association studies (GWAS) of psychiatric disorders identified various genes involved in synaptic functioning^4,16–18^, and gene expression studies identified differential expression patterns in synapse related genes in these disorders^19^.

While both neuroimaging and genetic studies each have pointed to synaptic alterations in psychiatric disorders, only a few have specifically tested this hypothesis in an integrated imaging-genetics framework. A few studies have explored the genetic architecture of functional brain connectivity^20–24^, and studies assessing polygenic risk scores have indicated links between psychiatric disorders and abnormal brain connectivity^25,26^. Previous studies also illustrated genetic correlation between various brain imaging phenotypes and psychiatric disorders that confirm a large degree of shared effect sizes across single nucleotide polymorphisms (SNPs)^27–29^. However, we still lack a concise map of the overlap in genetic architecture between psychiatric disorders and the brain functional connectome.

Recent evidence from anatomical imaging suggests a distributed nature of genetic effects on the brain, calling for tools that take a multivariate approach to imaging genetics, beyond univariate genome-wide association studies of single brain phenotypes^30^. We hypothesized that such distributed nature of genetic effects is also observable in the genetic architecture of functional imaging given the functional interplay of brain regions (nodes) in the connectome. A multivariate approach would perform better at capturing these distributed effects than conventional univariate GWASs^30^. Using data from the UK Biobank, we therefore deployed such approach to study the genetic architecture of functional brain connectivity – here defined as the correlation between time series data of large-scale brain network nodes^31,32^. Based on previous research pointing at dysconnectivity in psychiatric disorders, we expected that there is overlapping genetic architecture between the functional connectome and the disorders that can be captured using our multivariate approach. We therefore assessed genetic overlap between the connectome and eight major psychiatric disorders (ADHD, AN, ANX, ASD, BIP, MDD, PTSD and SCZ; Suppl. Table 1).

## Results

### Multivariate and univariate genome-wide association studies of the connectome

We performed two multivariate GWAS using the Multivariate Omnibus Statistical Test (MOSTest)^30^, which exploits the shared signal between related traits to discover genetic variants associated with the traits in a multivariate fashion. We performed one MOSTest analysis based on connectivity of 210 connections between 21 large-scale brain network nodes derived from an independent component analysis (ICA), and one based on signal variance across the duration of the functional scan in the respective 21 nodes. The two measures (connectivity and node variance) are mathematically related, yet they reflect different properties of functional brain connectivity, specifically properties of within large-scale brain network connectomics (node variance; referred to as node level) and between network connectomics (connectivity; referred to as edge level). Both measures have previously been found to be altered in psychiatric disorders^33^. The main analysis included data from 30,701 white British individuals aged 45-82 years (52.8% females) and replication analysis included an independent sample of 8954 individuals (42% white British) aged 45-83 (52.9% female). The Miami plots in Figure 1A illustrate the multivariate genetic associations calculated using MOSTest (N=30,701), and for comparison the associations identified using the traditional min-p approach, which takes the smallest p-value across univariate GWASs. Supplementary Figure 1 depicts corresponding QQ-Plots. MOSTest identified 15 genetic loci significantly (P < 5e-8) associated with functional brain connectivity (FC) and 5 loci significantly associated with node variance (Suppl. Table 2), whereas the min-p approach only identified 2 loci for FC and 3 loci for node variance. Four of the five loci identified for node variance were also present for FC, in line with the phenotypic relationship between the two. Replication analysis in the independent replication sample (N=8954) confirmed robustness of the main analysis, with 14 of 15 connectivity loci and all node variance loci replicating at the targeted multivariate replication rate of P < .05 (Figure 1C). Furthermore, to test whether the method by which we derived the functional brain measures affected the results, we supplemented our data-driven ICA approach with a separate region of interest (ROI) based pipeline, where we defined brain networks using an atlas of brain regions (see *Methods*). Results of these analyses indicate converging results despite an independent processing pipeline and network definition approach (Suppl. Fig. 2, Suppl. Table 3).

**Figure 1.**
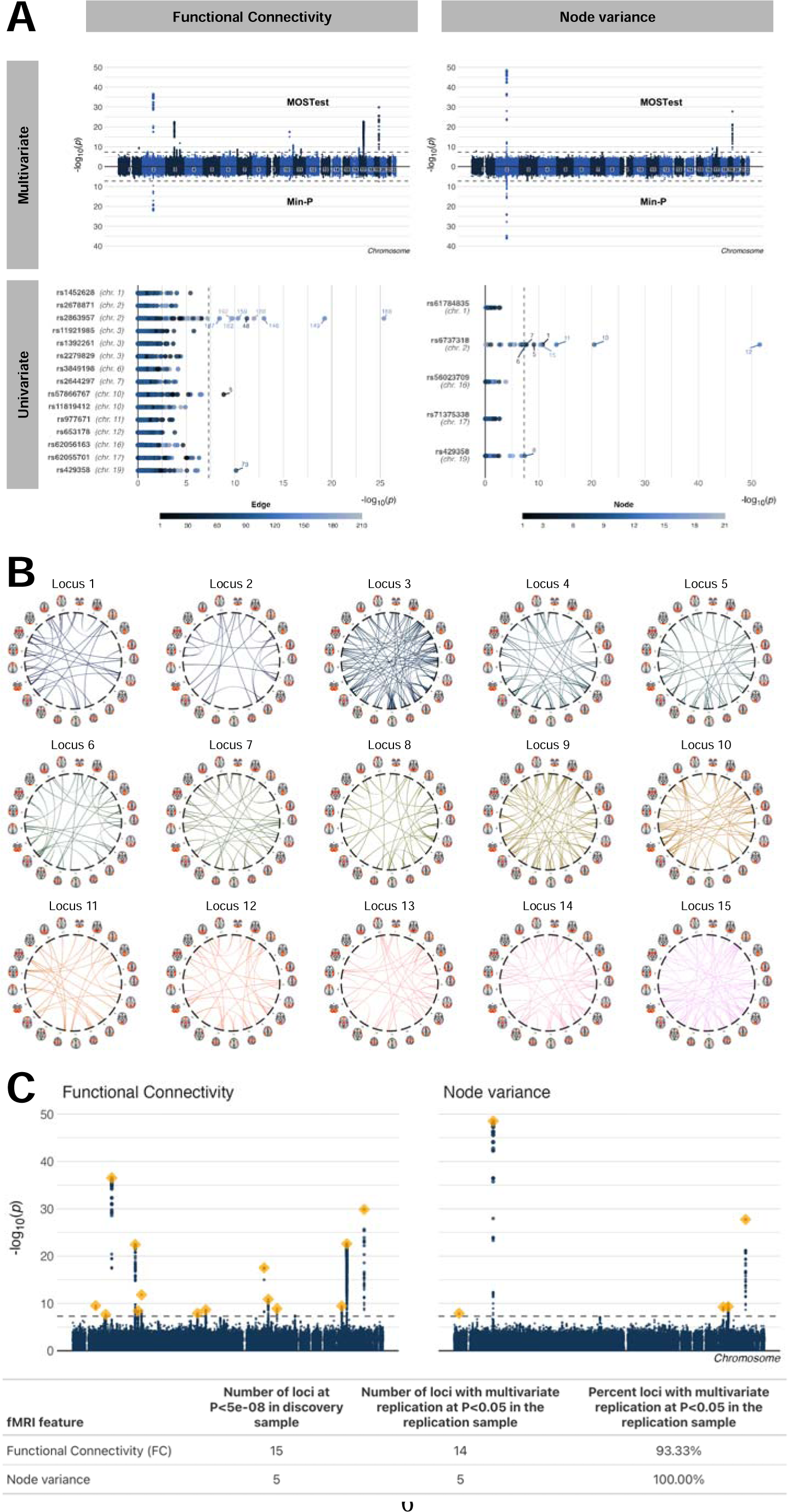
Multivariate and univariate architecture of the brain functional connectome highlight a distributed nature of effects across the brain. (A) The left column of the figure illustrates the results for functional brain connectivity, the right column for node variance. The first row shows Miami plots with the multivariate GWAS results from the MOSTest approach in the top, and the results from the traditional min-p approach at the bottom. The second row shows for each locus identified by MOSTest, the univariate p-values of the lead SNP in each locus. A majority of loci identified by the multivariate approach were not detected via the univariate approach. P-values are two-sided. (B) For each of the genome-wide significant loci underlying functional brain connectivity identified using the multivariate MOSTest approach, this panel shows nominally significant (P<0.05) connections from corresponding univariate statistics. These figures show differential patterns of regional SNP effects and highlight the distributed nature of the genetic effects on connectivity. (C) Results from replication analysis. The Manhattan plots depict the MOSTest summary statistics from Panel A and the loci replicated at nominal P < .05 in the independent replication sample are shown in orange. The table specifies corresponding multivariate replication numbers.

The bottom row in Figure 1A shows individual univariate p-values for the MOSTest-discovered loci, illustrating that the univariate approach is only good at capturing strong effects (e.g. locus 3 for FC), yet fails to discover loci with enriched signal across brain phenotypes. This also indicates that signal captured by the min-p approach reflects mostly the effect of individual phenotypes, rather than the combined signal as captured by MOSTest. Figure 1B further illustrates the distributed nature of effects across the brain, where a given locus shows differential patterns of regional SNP effects. Finally, genetic correlation analysis of univariate node variance GWAS (Suppl. Fig. 3) illustrated strong genetic correlations between different brain network nodes, largely in line with the phenotypic correlations observed when correlating the fMRI time series, adding further support to a distributed nature of effects in fMRI-based connectomics.

To complement the multivariate stream, we further analyzed the univariate GWAS for each connection in the full brain network and for each node variance separately, investigating whether the identified architecture from MOSTest is largely determined by a few network nodes or connections contributing with prominent signal or if it is determined by many nodes or connections, each contributing with subtle signal. Figure 2 depicts the SNP-based heritability for each connection (panel A) and for each node (panel B). SNP-based heritability ranged from 0.14% to 10.58% for brain connectivity (for 7 connections it could not be computed) and 137 out of 210 connections had a heritability above 1.96 times its standard error, indicating genetic signal^34^. The connection with the highest heritability was the connection between nodes reflecting activity in the prefrontal cortex (network 16) and the frontal network (network 14). For node variance, SNP-based heritability ranged from 3.92% to 13.64% with all nodes above 1.96 times their standard error, and highest heritability observed for node 9 (temporo-parietal network). Univariate analysis revealed no significant loci for any of the nodes or edges when controlling for the total number of edges or nodes through Bonferroni correction. The number of significant loci for the multivariate stream compared to the univariate stream adds further support that the genetic signal is distributed across the brain functional connectome, allowing us to capitalize on the shared signal for loci discovery.

**Figure 2.**
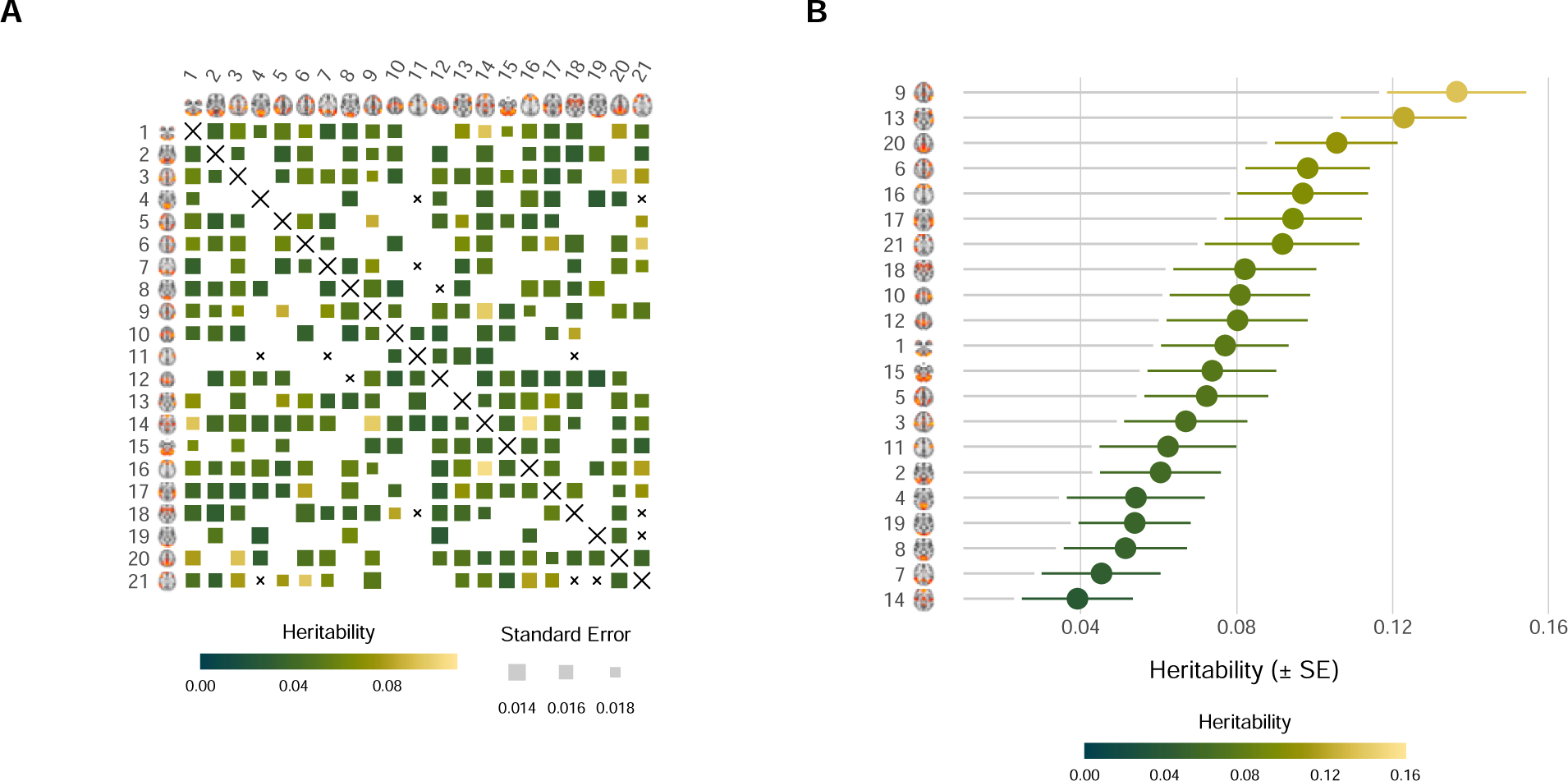
Heritability across edges and nodes. (A) SNP-based heritability (h2) for the 210 edges based on N=30,701 individuals. Upper half and lower halves of the figure are identical. Dark green indicates lowest heritability, bright yellow indicates highest heritability. Sizes reflect the standard error. Edges that did not survive heritability threshold are greyed out. Edges for which heritability could not be calculated are marked with a cross. (B) Point estimate of h2 (± SE) for the 21 nodes. Color scheme follows panel A. Data from N=30,701 subjects was used for estimates, error bars reflect the standard error

### Genetic overlap between connectome and psychiatric disorders

Next, using conjunctional FDR analysis^35^, we tested for overlap between the two MOSTest-derived genetic profiles (functional connectivity and node variance) with eight major psychiatric disorders, specifically Attention-Deficit Hyperactivity Disorder (ADHD)^36^, anorexia nervosa (AN)^37^, anxiety disorder (ANX)^38^, autism spectrum disorder (ASD)^39^, bipolar disorder (BIP)^40^, major depression (MD)^41^, Post-Traumatic Stress Disorder (PTSD)^42^, and schizophrenia (SCZ)^43^. Conjunctional FDR leverages pleiotropic enrichment (statistical pleiotropy^44^) between two phenotypes to identify genetic loci jointly associated with them. As shown in Figure 3, we found shared loci for seven of the eight disorders, namely for ADHD, AN, ANX, ASD, MDD, BIP and SCZ. By far the largest number of shared loci was implicated for SCZ (43 for FC, 22 for node variance). We found 6 loci for FC and 1 locus for node variance in ADHD, 9 loci for FC and 2 loci for node variance in BIP, and 4 loci for FC and 3 loci for node variance in ASD. Additionally, we found 1 shared locus between FC and MDD, 1 shared locus between FC and AN, and 1 shared locus between node variance and ANX. We did not find any shared loci between either FC or node variance and PTSD. Supplementary Figure 4 depicts quantile-quantile plots for all genetic overlap analyses. Additional sensitivity analyses using a more stringent FDR threshold confirmed largest overlap for SCZ with FC among the traits (Suppl. Table 4). Analysis with a negative control trait (vitamin D levels: N = 79,366)^45^ yielded no significant overlap for node variance and two loci for connectivity (Suppl. Fig. 5). Finally, re-analysis with the ROI-based pipeline confirmed genetic overlap identified in the ICA-based analysis, again with most loci implicated for SCZ (Suppl. Fig. 6).

**Figure 3.**
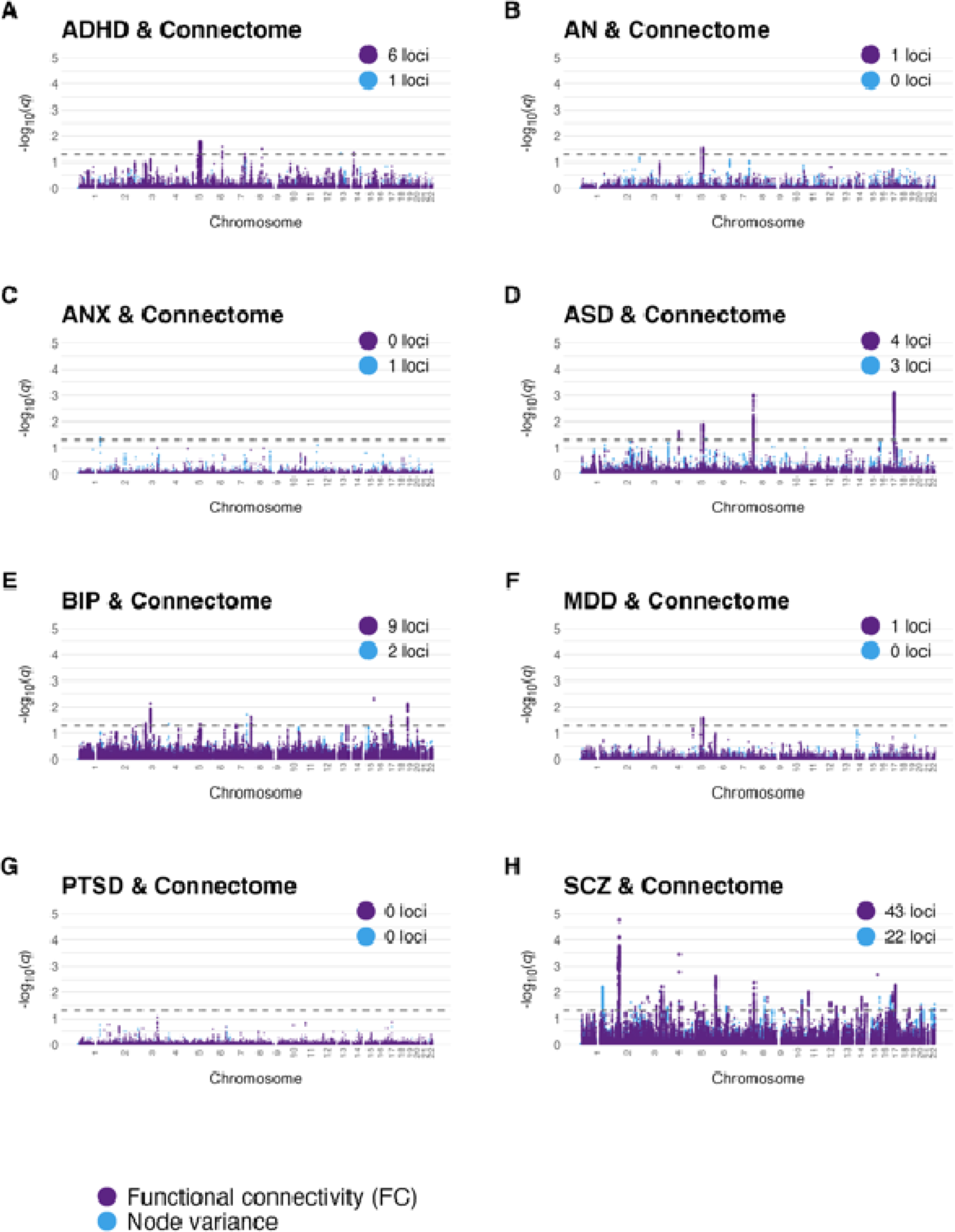
Manhattan plots illustrating genetic overlap between disorders and the multivariate functional brain phenotypes. Each panel (A-H) shows the association each psychiatric condition. for Association strength per locus is depicted as q-value from the conjunctional FDR analysis^46^. Values for FC and node variance are shown in the same figure with separate colors.

Using Functional Mapping and Annotation of GWAS (FUMA)^47^, we mapped the loci shared between the connectome and the psychiatric disorders to 180 genes, listed in Suppl. Table 5 (for comparison to ROI-based analysis, see Suppl. Table 6). We tested for enrichment for biological processes (GO) and identified 125 significant associations, many of which are relevant to neural system development and functioning (Suppl. Fig. 7). Using *SynGO*^48^, we linked 23 of the 180 genes to synapse functioning (Suppl. Table 7). For example, one of the loci shared between SCZ and FC was mapped to BDNF, which is a major regulator of synaptic transmission and synaptic plasticity^49^. Another example is NRXN1, found also for SCZ and FC, which is known for its role in the formation of synaptic contacts^50^. Utilizing the pathway browser on the identified gene sets^51^, we also found that the mapped genes were involved in cell signaling and signal transduction, more specifically protein-protein interactions at the synapses, WNT and NTRK signaling, but also a number of other biological processes such as chromosome maintenance and mitosis (Suppl. Fig. 8 and Suppl. Table 8).

In addition to the conjunctional FDR analyses, we also calculated genetic correlation between each connection or node surviving our pre-defined threshold of 1.96 times its SE and the eight psychiatric disorders, allowing us to compare the multivariate findings to results from a univariate approach. Figure 4A illustrates that genetic correlation was generally low for the connectome and only one connection survived after correcting for all eight disorders and all connections, specifically a connection between the right ventral (network 21) and the prefrontal network (network 16) was significantly associated with BIP (r_g_ = −0.25, p_BONF_ = 0.0006). When only correcting for the number of connections but not for the number of disorders, we found an additional significant association, which was the link between the auditory (network 17) and the subcortical (network 18) node which correlated with SCZ (r_g_ = 0.25, p_BONF_ = 0.0137). For node variance, we found two significant associations when correcting for all disorders and nodes. Specifically, variance in the temporo-parietal network (network 9) was significantly correlated with both, SCZ (r_g_ = 0.22, p_BONF_ = 3.9e-6) and BIP (r_g_ = 0.17, p_BONF_ = 0.03).

**Figure 4.**
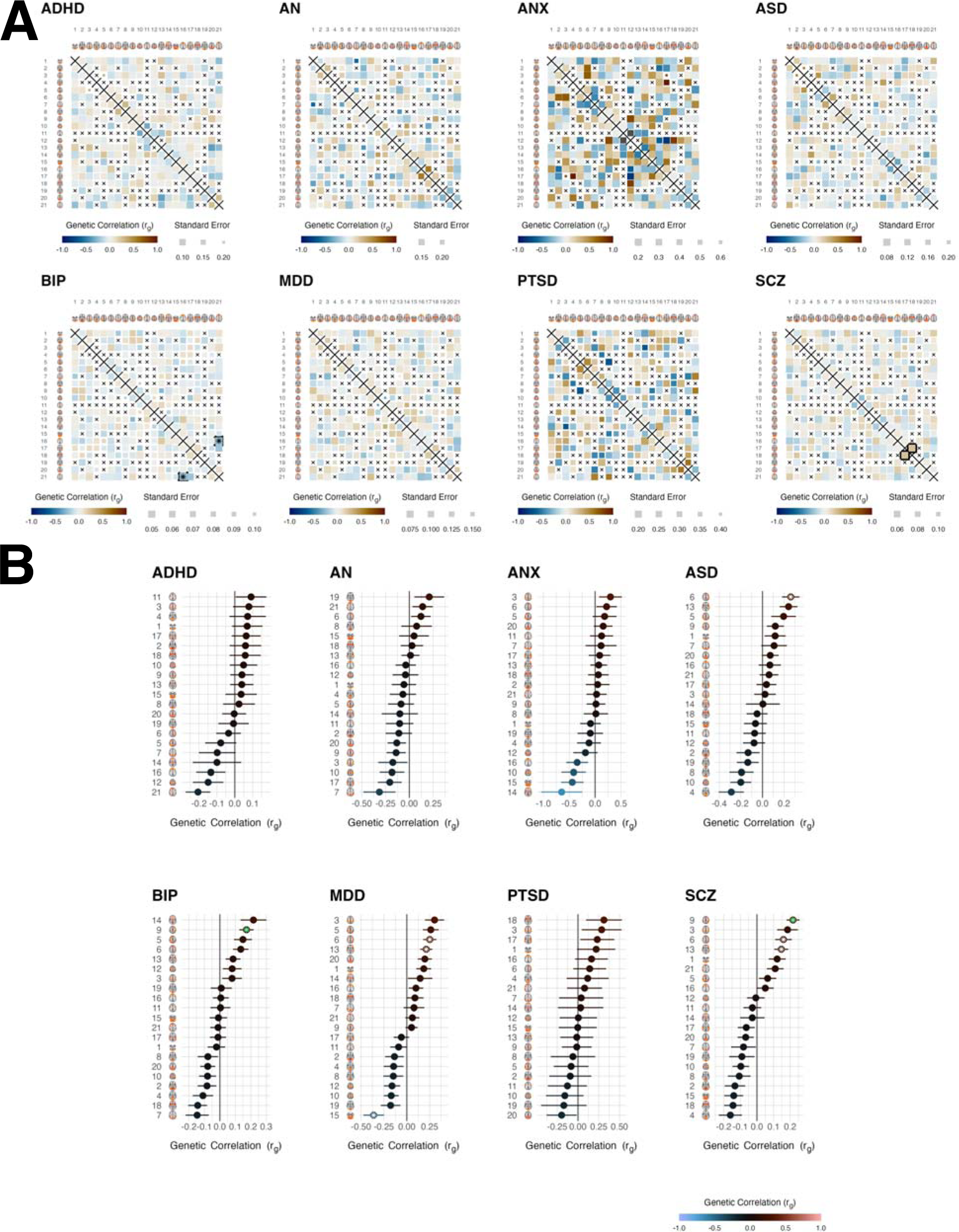
Genetic correlation between the connectome and psychiatric disorders. (A) The tiles show the genetic correlations between each edge of the whole brain network and a given psychiatric disorder. Size of the tile represents the standard error. Edges with a heritability below 1.96 its standard error were not considered in the analysis and marked with a black cross. Among all disorders, only one edge marked with a black border was significant for SCZ after correcting for the number of edges (210), whereas none was significant when correcting for the number of edges and the number of disorders. Upper and lower half of each matrix are identical. Sample sizes underlying the summary statistics used for genetic correlation analysis are provided in Supplementary Table 1. (B) Genetic correlation (± SE) analysis at the node level. Significant genetic correlations within a psychiatric disorder are indicated with a white asterisk when correcting for the number of nodes, whereas a green asterisk indicates significance when correcting for both, the number of nodes and the number of disorders. The error bars reflect the standard error. The latter stringent correction was surpassed for variance of the temporo-parietal node and SCZ.

## Discussion

Taken together, our study provided insight into the shared genetic architecture between measures of the brain functional connectome and common psychiatric disorders. Deploying multivariate genetic analyses of fMRI data from more than 30,000 individuals allowed us to capitalize on the distributed nature of genetic variation across the interconnected whole brain network to discover novel connectome-associated variants beyond what can be discovered using standard univariate approaches. Our analyses pinpointed a number of gene variants overlapping between the connectome and psychiatric disorders, where several of the corresponding mapped genes are known for their involvement with synapse formation and functioning.

We used two measures of the brain functional connectome – the 210 correlations of brain signal from 21 nodes as measures of functional brain connectivity as well as signal variation across time of these 21 network nodes. Given the interconnectedness of the connectome, we hypothesized that many connections or nodes would have overlapping genetic signatures. Indeed, our results illustrate that the genetic architecture of brain function is distributed across the brain. Our deployed multivariate approach successfully leveraged this pleiotropy for discovery, revealing a variety of genetic effects that would not have been discovered with the standard univariate GWAS approach, including the commonly used min-p approach, which identifies the minimum p-value across univariate GWASs. We observed that the significant lead SNPs from MOSTest were often not significantly associated with the univariate measure. This demonstrates that using multivariate genetic analysis can be valuable to complement the univariate approach in settings like brain imaging where the signal is largely distributed.

From our multivariate signatures of the connectome, we were able to identify a number of shared loci with psychiatric disorders through conjunctional FDR analysis. The strongest overlap was implied for schizophrenia yet all other psychiatric disorders apart from PTSD showed some degree of overlap as well, in particular with connectivity. Identification of overlap to some degree depends on statistical power (overall heritability, sample size, quality of phenotyping, heterogeneity across contributing cohorts, among others), which may for example explain the lack of findings for PTSD. It is important to note that sample size alone cannot explain the observed differences in overlapping loci. The bipolar disorder and schizophrenia GWASs have similar sample sizes yet we discovered many more loci for the latter. Likewise, our negative control analysis of a well powered trait yielded only two overlapping loci for connectivity and no significant locus for node variance. How the comparison between all disorders will look at similar sample size remains to be investigated.

Several synapse-related genes were among the overlapping genes, including some involved in the neurodevelopmental formation of synapses. This is particularly intriguing given that many psychiatric disorders are conceptualized as neurodevelopmental disorders even if they are typically diagnosed in adulthood. Further, many disorders are conceptualized as disorders of brain dysconnectivity, as initially proposed for schizophrenia^52^. This is now established across various disorders^53^ and our results provide further evidence from the genetics end.

While multivariate analysis enabled us to boost discovery across the imaging phenotypes, its multivariate nature limits interpretation of single features, such as for example biological interpretation of specific brain networks. We thus complemented our multivariate analyses with univariate analyses and showed a map of genetic correlations between connectivity, node variance and psychiatric disorders. Univariate correlations were overall weak and only a few edges or nodes were significant after correcting for multiple testing. A particular pronounced univariate association was found for the temporo-parietal network, which was genetically correlated with schizophrenia (p_BONF_ = 3.9e-6) and bipolar disorder (p_BONF_ = 0.03). Strikingly, in a previous fMRI study this temporo-parietal network was found to be consistently altered in schizophrenia across five different fMRI tasks, with bipolar disorder clustering between schizophrenia and healthy controls in three out of five tasks^54^. Nonetheless, given the overall weak univariate associations and the large amount of univariate analyses performed, caution is warranted when interpreting potential disorder-specific patterns. Lack of genetic correlating does not necessarily indicate lack of polygenic overlap, as shown previously by multiple methods, including cross-trait MiXeR analysis^55^ and LAVA^56^. It is well possible that a variety of SNPs with opposing effect directions cancel each other out, which would result in low genetic correlation despite genetic overlap. The finding that genetic correlations from univariate analyses are relatively weak despite significant genetic overlap between our multivariate GWAS and several psychiatric disorders provides further evidence that multivariate analysis is an important method to dissect complex interactions in psychiatric genetics.

We here provided results from analyses at the edge level (functional connectivity) and node level (node variance). The latter showed larger heritability and larger effect sizes than functional connectivity. This may be partly explained by the granularity of the connectivity measure, or a better representation of the nodes across individuals compared to the potentially highly individualized network configurations^57,58^. At the phenotypic level, node variance has been associated with psychiatric disorders, with effect sizes comparable to connectivity^33,59,60^. Given that our genetic analyses often imply similar genes for node-level and connectivity-level, the underlying sources may align despite differences in current association effect sizes.

Some aspects are relevant for interpreting the current findings. First, MOSTest is to some degree dependent on granularity as also previously shown^61^ which may explain why MOSTest identified more loci for functional connectivity than for node variance. More research using different approaches to network definition may yield further discoveries, however, our comparison of the genetic architecture of ICA-based and ROI-based networks also indicated large overlap, supporting robustness of the current findings. While the granularity of the ICA-based and ROI-based networks were kept similar in this analysis, there is still substantial difference in their derivation, and thus it is remarkable that we reproduce our ICA-based genetic findings with the ROI-based approach. Specifically, the spatial pattern of ICA-based networks are data-driven, and connectivity is determined by means of regularized partial correlations. In contrast, ROI-based networks are spatially defined with an atlas and connectivity is estimated via full correlations. We here analysed large-scale brain networks. Technically, MOSTest is capable to scale to much large numbers of input features^62^ and thus analyses with more fine-grained parcellations or even at the vertex or voxel level might be possible. Such scaling would need to be performed under careful observation of noise structures in the functional imaging data to ensure that the potential benefit of more features is not diminished by added noise. Second, lack of effect directionality is a limitation of the multivariate analysis, which is why we provided univariate analyses alongside. Furthermore, several post-GWAS analyses such as for example the conjunctional FDR framework do not require effect direction and can thus be performed with the resulting multivariate statistics. We believe the strengths of the multivariate approach outweigh the limitations, and a tandem approach with both multivariate and univariate methods optimizes utility of this method. Finally, with conjunctional FDR analysis it can happen that some discoveries are novel, meaning they are missed by standard GWAS due to lack of power or excessive burden of multiple testing. However, it is expected that conjunctional FDR loci will be discovered by these standard GWAS methodology once sample sizes increase further. For example, two loci recently discovered in a GWAS on ADHD^36^ where already discovered earlier using a conjunctional FDR analysis with educational attainment^63^.

In conclusion, we here revealed a distributed nature of genetic effects on brain function and integration, and identified a number of genetic loci associated with key properties of the brain functional connectome. Further, we revealed a large degree of genetic overlap between multivariate measures of the brain functional connectome and a number of psychiatric disorders with genes pointing at synaptic plasticity. This may help further disentangle the complex biological underpinnings of psychiatric disorders and provide a bridge between functional connectivity alterations and genetic variations in patients. There is a need for follow-up experimental studies building on the discovered loci to disentangle the biological mechanisms.

## Methods

### Sample and Exclusion Criteria

We accessed resting state fMRI data from the UK Biobank^64^, a large-scale resource of imaging, genetics, and other biological and psychological data (access with permission no. 27412). All participants provided signed informed consent before inclusion in the study. The UK Biobank was approved by the National Health Service National Research Ethics Service (ref. 11/NW/0382).We selected data from individuals with White British ancestry, identified based on the genetic clustering performed by the UK Biobank team^65^. Data of all eligible participants were included for the main analysis in November 2020 and we did not exclude individuals based on a diagnosis. The resulting sample comprised data of 30,701 individuals with a mean age of 64.24 years (SD: 7.50, range: 45-82; 52.8% females). In total 2654 individuals (8.62%) had a neurological or psychiatric diagnosis (excluding encephalitis). Additional data became available afterwards and was partly used for replication (see Replication section).

### Image Acquisition and Preprocessing

Data had been acquired by the UK Biobank study team^64^. The fMRI images were collected on four identical 3T Siemens Magnetom Skyra scanners in the UK with a 32 channel head coil (Siemens Healthcare GmbH, Erlangen, Germany). Data was recorded using a gradient-echo echo planar imaging sequence with x8 multislice acceleration (TR: 0.735s, TE: 39ms, FOV: 88×88×64 matrix, FA: 52°) with a voxel size of 2.4×2.4×2.4mm. One fMRI sequence took approximately 6 minutes. The protocol further included T1 imaging, acquired using a MPRAGE sequence with in-plane acceleration (iPAT) of 2 (resolution: 1mm^3^, FOV: 208×256×256 matrix).

Data had been preprocessed by the UK Biobank study team as described in Alfaro-Almagro *et al.*^66^. Briefly, their preprocessing used the FSL pipeline^67,68^, which included motion correction using MC-FLIRT (Jenkinson, Bannister, Brady, & Smith, 2002), grand-mean intensity normalization, high-pass filtering through Gaussian-weighted least-squares straight line fitting, EPI unwarping and GDC unwarping. Structured artifacts were removed using ICA and FIX^69,70^, where the FIX classifier was hand-trained on 40 UK Biobank datasets. According to Alfaro-Almagro *et al.* only 1% of variance in a scan is due to head motion following motion correction and FIX^66^. The final step was a group ICA using MELODIC^71^ which decomposed the data using independent component analysis into 25 components. The spatial profiles of the components can be reviewed in a navigable visualization tool available at https://www.fmrib.ox.ac.uk/ukbiobank/group_means/rfMRI_ICA_d25.html.

We retrieved individual level time series data for each subject and component (output from dual regression at model order 25). We computed functional brain networks using the FSLNets toolbox^72^. First, we regressed the time series of four noise components from the time series of the remaining 21 components and subsequently removed those four components. Suppl. Fig. 9 depicts maps for each of the 21 components. We estimated functional connectivity (FC) as the regularized partial correlations of the component time series, implementing an approach developed by Ledoit & Wolf (2012) which performs an automated adjustment of the shrinkage parameter lambda, as implemented in our earlier work^54^. As the last step, we regressed age, age^2^, sex, scanner, motion, signal-to-noise ratio (SNR), and the first 20 genetic principal components from the individual connection strengths, residualizing each edge (210 in total) of the partial correlation matrix. In addition to functional brain connectivity, we also performed an analysis of the variance in signal amplitude of the 21 components^67^, and performed the same residualisation in this node-level analysis as described above for the edge level.

To test if our results were largely dependent on the pipelines used to define brain networks, we complemented our main data-driven ICA approach with a region-of-interest (ROI) approach using the Schaefer parcellation with 1000 parcels. For this pipeline we accessed FEAT^73^ processed folders from the UK Biobank and registered all images to standard MNI space. For each Schaefer-defined ROI there exists a mapping to the 17 large-scale brain networks defined by Yeo et al (2011). To achieve comparability to our main ICA-based analysis which comprises 21 network nodes, we averaged the time series of all Schaefer-defined ROIs corresponding to each Yeo-defined network, yielding ROI-based networks with 17 nodes. Following the standard procedure for ROI-based brain networks, we defined these as the Pearson correlation of the 17 nodal time series. Furthermore, we derived node variance of these 17 nodes. The resulting functional brain connectivity as well as node variances went into the same genetic analyses as performed in the main ICA-based analysis workflow.

### Genetic data and QC

We accessed UKB v3 imputed data^64^. The data acquisition and preprocessing pipeline is described in Bycroft *et al.*^64^. We applied standard quality control procedures to this data and removed SNPs with a minor allele frequency below 0.001, SNPs missing in more than 5% of individuals, SNPs with an imputation quality below 0.5, and SNPs failing the Hardy-Weinberg equilibrium test at P<1e-9.

### Univariate and Multivariate Genome-Wide Analysis

We performed multivariate and univariate GWAS using the Multivariate Omnibus Statistical Test (MOSTest)^30^. MOSTest takes as input all univariate test statistics (*z*-scores) for each SNP, as obtained through standard association testing with each pre-residualized phenotype, and compares this to test statistics obtained following a single random permutation of the genotype vector. A multivariate test statistic is then calculated from this comparison as the Mahalonobis norm, with the probability of the observed test-statistic being derived from a Chi-square distribution. Further details of the method are described in Van der Meer *et al.*^30^. MOSTest returns a multivariate test statistic, where in contrast to classical univariate GWAS that link a given SNP with a single phenotype, for each SNP the multivariate association across all included phenotypes is provided. This allowed us to retrieve one multivariate summary statistic for functional brain connectivity (edge level), and one for node variance (node level). In addition, we retrieved classical univariate summary statistics for follow-up analyses.

### Summary Statistics for Psychiatric Disorders

We accessed publicly available summary statistics for Attention-Deficit Hyperactivity Disorder (ADHD)^36^, anorexia nervosa (AN)^37^, anxiety disorder (ANX)^38^, autism spectrum disorder (ASD)^39^, bipolar disorder (BIP)^40^, major depression (MD)^41^, Post-Traumatic Stress Disorder (PTSD)^42^, and schizophrenia (SCZ)^43^. For details, see Suppl. Table 1. We used vitamin D^45^ as a negative control phenotype because it is well powered (N = 79,366), has heritability comparable to psychiatric disorders (h2_twin_∼0.6) and is not genetically correlated with the included psychiatric disorders (all P>.05). We performed a GWAS using plink 2.0^74^ on 79,366 participants in the UK Biobank not included in the main analysis.

### Pleiotropy-Informed Conjunctional False Discovery Rate

Due to the complex and polygenic architecture of our brain phenotypes, we utilized pleiotropy-informed conjunctional false discovery rate as implemented in the pleioFDR toolbox^46^. The conjunctional FDR analysis identifies shared genomic loci between two traits regardless of effect directionality and effect size, making it ideally suited to compare a multivariate summary statistic from MOSTest (here: FC and variance) against the summary statistics of a given disorder (here: SCZ, BD, MDD, ASD, ADHD, ANX, PTSD, AN).

### Linkage Disequilibrium Score Regression

For the univariate summary statistics, we estimated partitioned heritability^75^ and genetic correlation with LD-score regression using the LDSC tool^76^. We also estimated genetic correlation between each edge and variance across time in each node with the eight psychiatric disorders using cross-trait LDSC^76–78^. Of note, genetic correlations require effect directions and are thus not applicable to the multivariate summary statistics derived from MOSTest. We therefore used genetic correlations in connection with univariate statistics as a complement to the multivariate pipeline.

### Gene Mapping and Annotation

We used the Functional Mapping and Annotation (FUMA version v1.3.6a) tool to map loci derived through conjunctional FDR analyses to genes and tested for significant enrichment of biological processes^47^. We then fed the genes identified through FUMA into the *SynGO* (v1.1) toolbox to map synaptic genes^48^, and the *reactome* (v78) toolbox to map the genes to a range of biological processes^51^.

### Statistics and Reproducibility

No statistical test was used to predetermine sample size. To validate the discovered loci of the functional brain measures, we performed a replication analysis of our two main MOSTest analyses on a dataset containing all subjects with available data (including those with non-White British ancestry) as well as a new batch of data (including White British) that arrived after we performed the main analyses. This resulted in a dataset containing 8954 individuals (mean age: 65.20 years, SD: 8.25, range: 45-83; 53.0% females). Of these, 5155 individuals were of non-White British ancestry and 3799 individuals were of White British ancestry. The replication sample included 1024 individuals (11.4%) with a diagnosed neurological or psychiatric diagnosis (excluding encephalitis). We processed this dataset in the same way as the data from the discovery sample. Multivariate discoveries require a special replication procedure to ensure that a locus in question is not only showing an association in an independent sample, but also that the multivariate pattern of that association is consistent between the discovery and the replication samples. Such procedure has been established in Loughnan et al.^79^ For a given SNP in the discovery set, the procedure provides a composite score (one value for each individual in the validation set) obtained as a weighted sum of individuals’ phenotypes, with weights derived from mass-univariate z-statistics from the discovery set. If a SNP association represents a real signal in the discovery set, we expect its composite score to be associated with the genotype in the replication sample at a nominal one-sided P<0.05, and to have a consistent effect direction. Mathematical formulation of the approach is provided in Loughnan et al.^79^

### Inclusion & Ethics

The samples used in this study comprised samples of varying ethnic backgrounds. While the main analyses unfortunately still incorporate only individuals with a White British ancestry, we used a more diverse sample to validate our in the replication analyses, to provide insights to make them relevant for application beyond this well-studied group of White British individuals. Given the limited availability of data from non-White individuals, the current work cannot rule out ethnicity biases, and replication in larger and more diverse samples is needed to further assess replication.

## Data Availability

Data used in this study are part of the publicly available UK Biobank initiative (https://www.ukbiobank.ac.uk/). Summary statistics for the disorders are publicly available through their respective consortia (Suppl. Table 1). The summary statistics for the multivariate analyses will be shared on GitHub (https://www.github.com/norment/open-science) upon acceptance.

## Code Availability

Code will be made publicly available via GitHub (https://www.github.com/norment/open-science) upon acceptance of the manuscript.

## Acknowledgements

The authors were funded by the Research Council of Norway (#276082 LifespanHealth, #323961 BRAINGAP, #223273 NORMENT, #283798 ERA-NET Neuron SYNSCHIZ, #249795, #298646, #300767), the South-East Norway Regional Health Authority (2019101, 2019107, and 2020086), and the European Research Council under the European Union’s Horizon2020 Research and Innovation program (ERC Starting Grant #802998), as well as the Horizon2020 Research and Innovation Action Grant CoMorMent (#847776). This research has been conducted using the UK Biobank Resource (access code 27412, https://www.ukbiobank.ac.uk/). This work was performed on the TSD (Tjenester for Sensitive Data) facilities, owned by the University of Oslo, operated and developed by the TSD service group at the University of Oslo, IT-Department (USIT). Computations were also performed on resources provided by UNINETT Sigma2 - the National Infrastructure for High Performance Computing and Data Storage in Norway.

## Author Contributions

D.R. and T.K. conceived the study; D.R. analyzed the data with contributions from T.K.; All authors contributed with conceptual input on methods and/or interpretation of results; D.R. and T.K. wrote the first draft of the paper and all authors contributed to the final manuscript.

