## Supplementary Figures for "Genetic overlap between multivariate measures of human functional brain connectivity and psychiatric disorders"

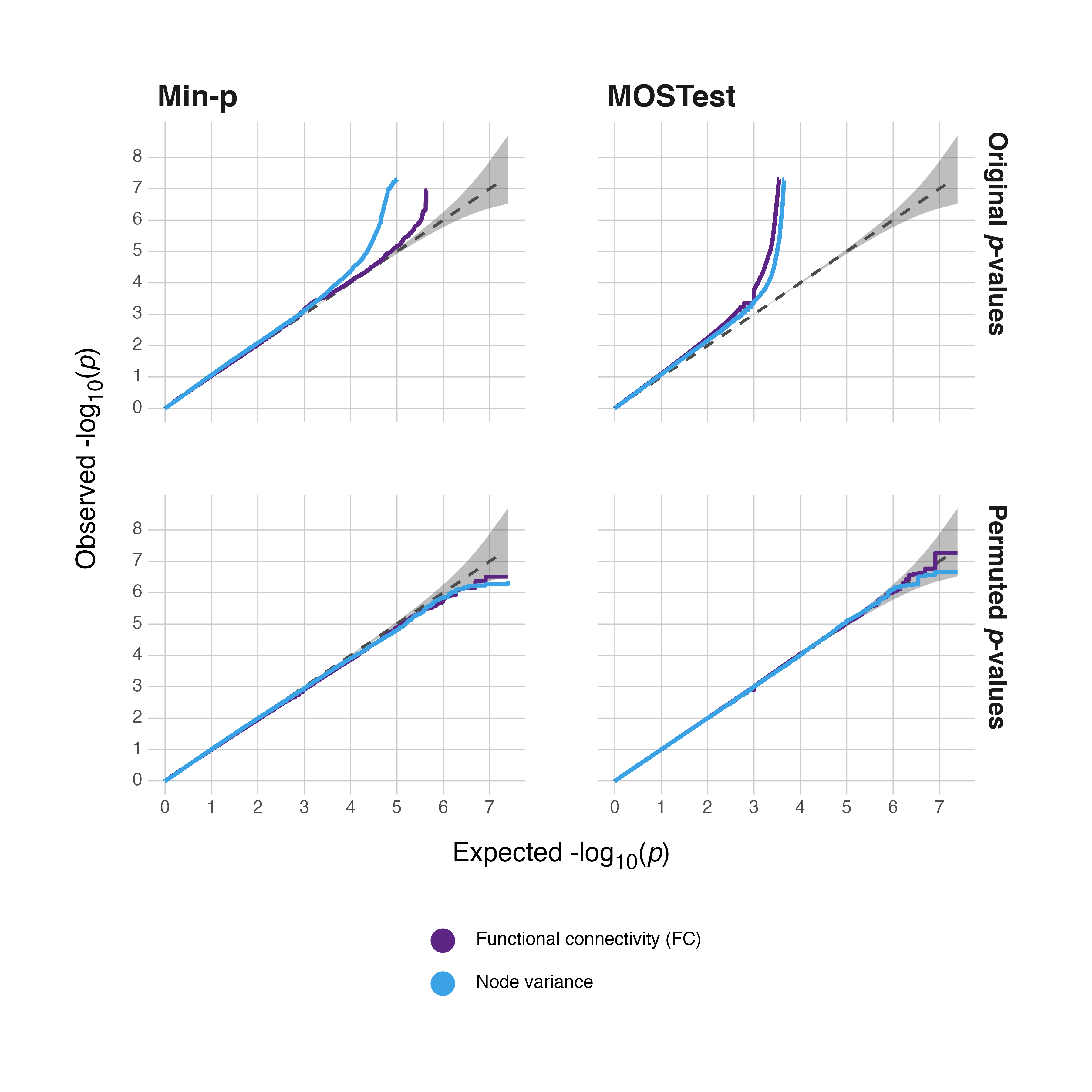

**Suppl. Fig 1.** **Quantile-Quantile plots corresponding to both the MOSTest and the min-p analysis.** The first column shows the QQ plots for MOSTest, the second column shows the QQ plots for the min-p approach. The first row shows the original p-values, the second row shows the permuted p-values. In both panels the shaded area represents the 95% CI. An upward deflection from the null-line indicates stronger genetic signal. As expected, the first row shows strong genetic signal whereas the permuted p-values do not reflect genetic signal. To increase legibility all data points larger than -log_10_(5e-8) are removed.

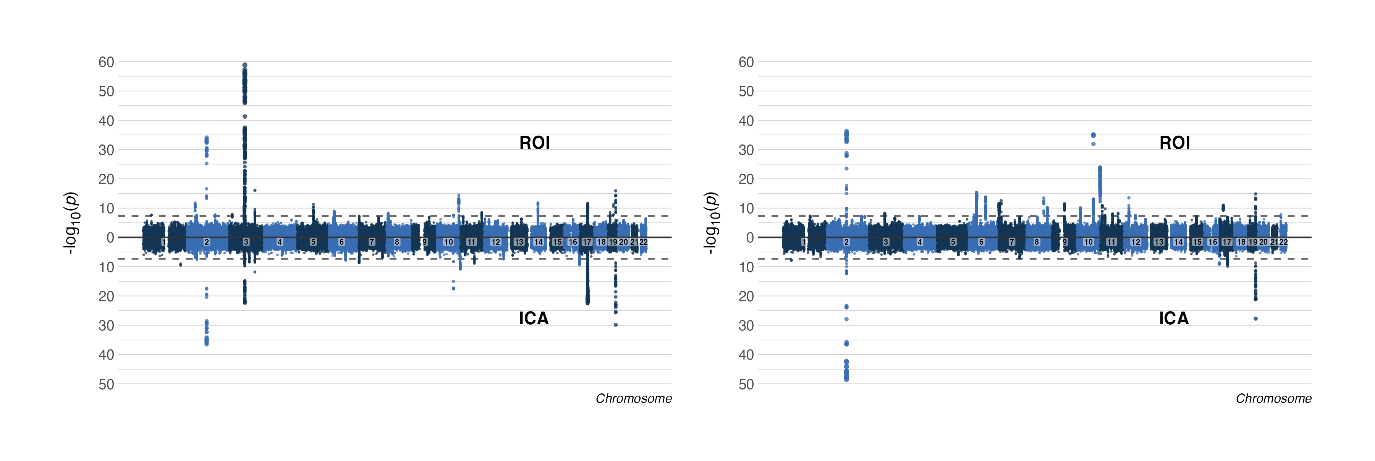

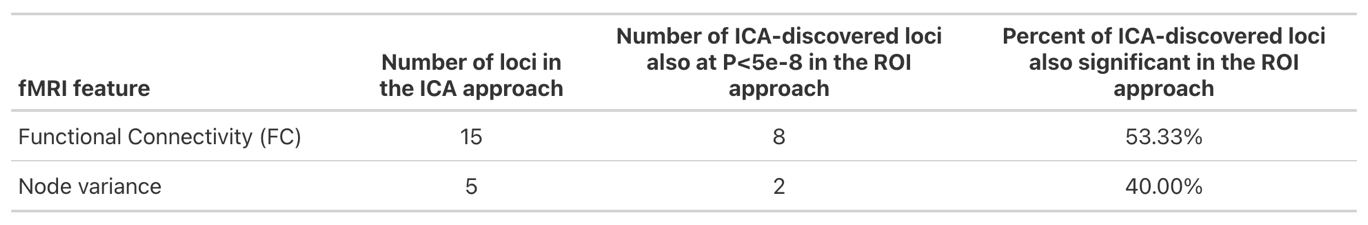

**Suppl. Fig 2. Miami plot comparing discovery between two different approaches for functional brain network definition.** The upper part of the Miami plots illustrate results based on a region-of-interest (ROI) approach, whereas the lower part illustrates results from the main ICA-based analysis. The left figure shows results for functional connectivity, the right figure shows results for node variance. The table specifies the number of loci identified in the ICA-based approach that were also identified in the main ROI-based analysis (i.e., have SNPs in the ROI-based analysis with P<5e-8). The results confirm strong overlap in discovery when using these two distinct approaches to derive the studied phenotypes.

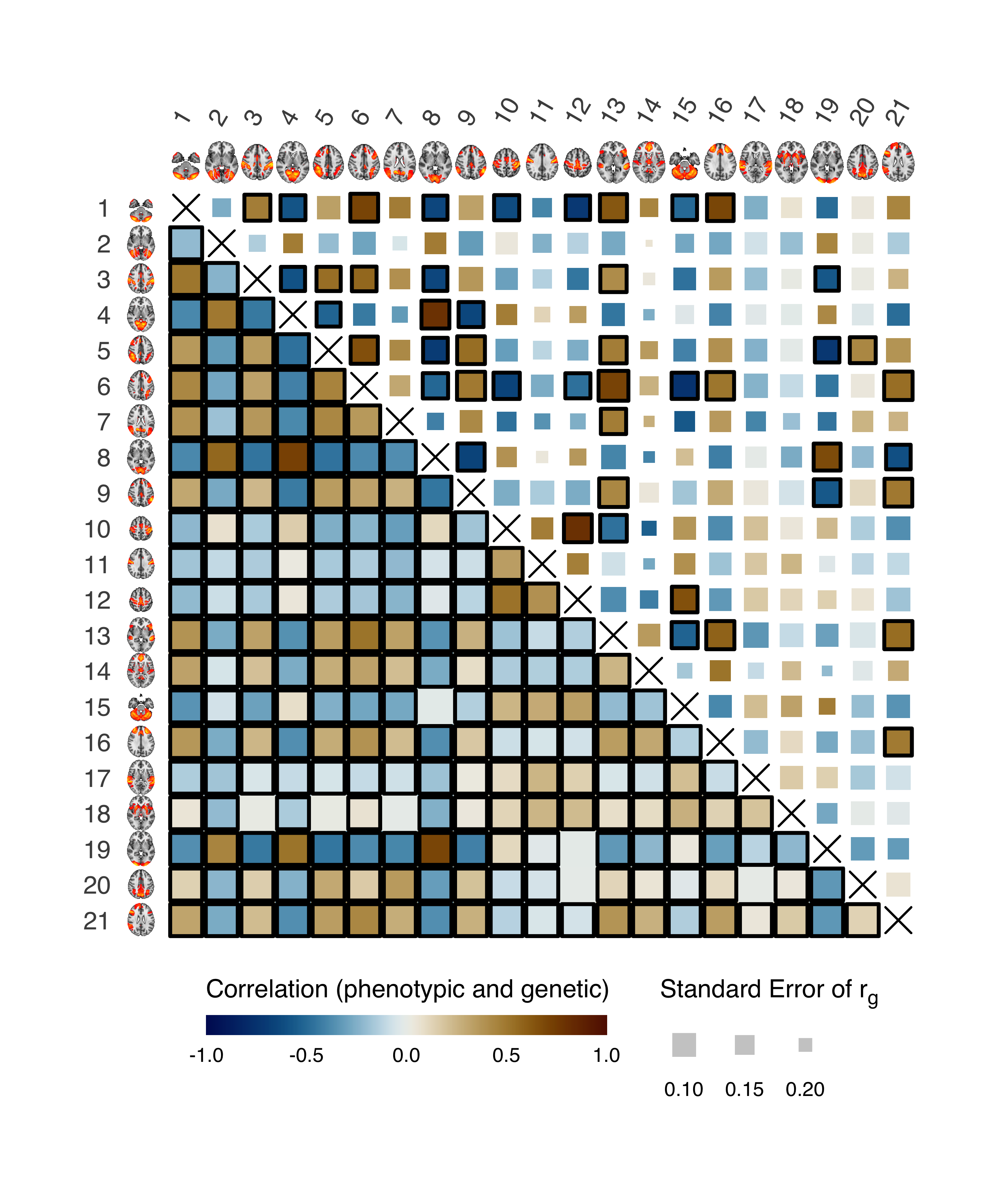

**Suppl. Figure 3.** **Genetic and phenotypic correlation between nodes.** This figure shows the genetic correlation between the nodes in the upper half (based on GWAS of node variance), and the phenotypic correlation in the lower half (based on correlations of the node time series). Standard error of the genetic correlation is indicated by the size of the tile. Significant genetic correlations are indicated by a black border. Phenotypic correlations were all significant except for 7 of them, while there were in total 46 significant genetic correlations (22%). For example, the strongest significant genetic correlation was between two nodes of the sensorimotor system (nodes 10 and 12, r_g_ = 0.8007, p_BONF_ = 4.0e-16). The second strongest genetic correlation was between different parts of the visual cortex (node 4 and 8, r_g_ = 0.7976, p_BONF_ = 2.1e-14).

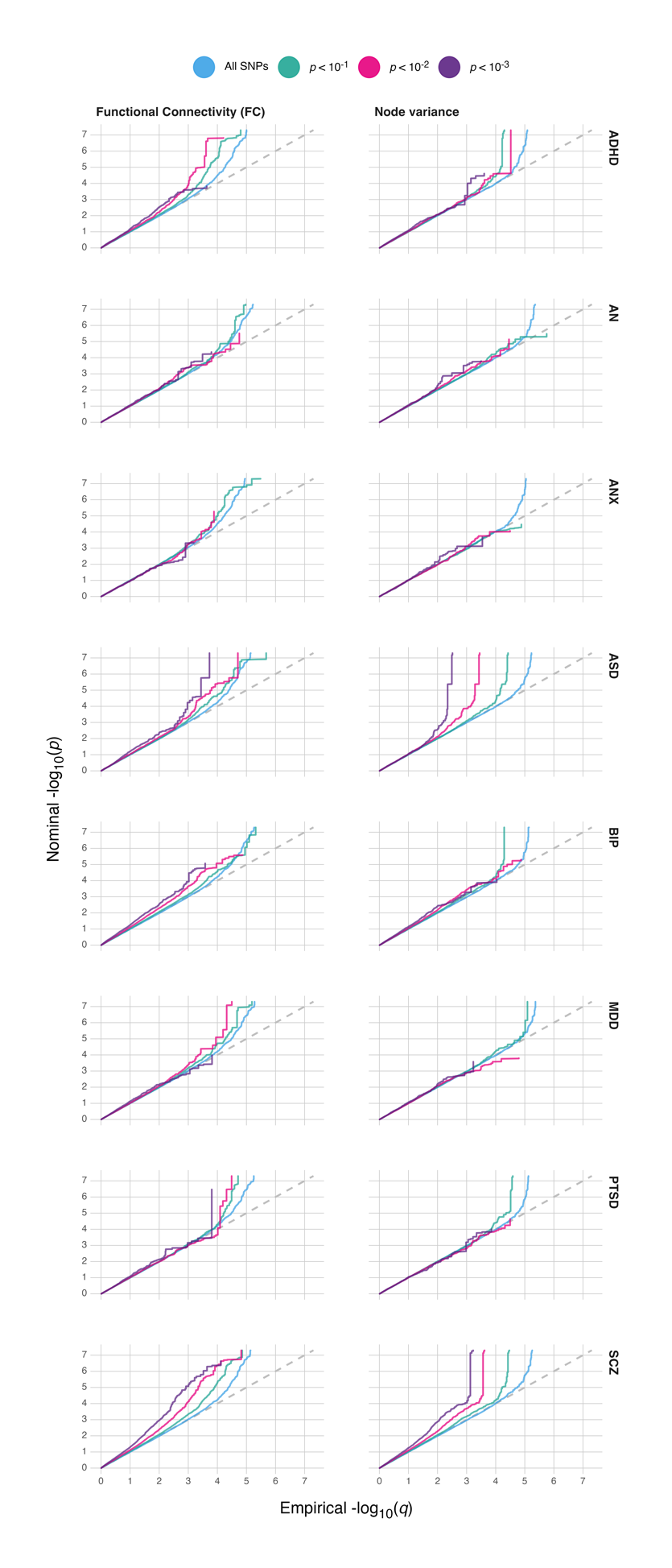

**Suppl. Fig 4.** **Quantile-Quantile plots for genetic overlap analysis.** The left column depicts FC, the right column depicts node variance and each row indicates overlap with one of the disorders. Each plot shows genetic signal when including all SNPs as well as three stratifications based on different significance thresholds. To increase legibility and comparability between the plots data outside the margin -log_10_(5e-8) is removed.

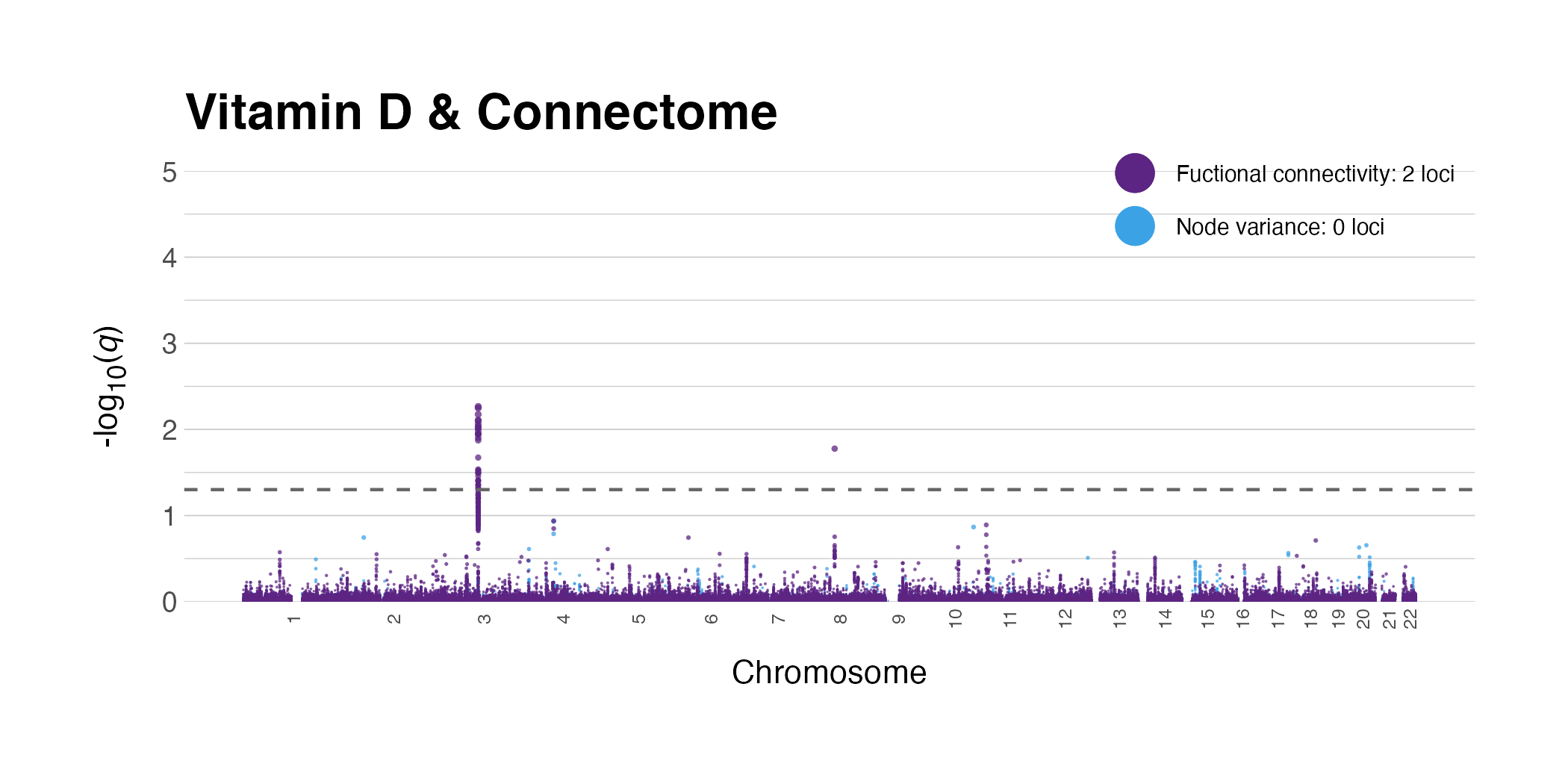

**Suppl. Fig 5. Negative control analysis.** Manhattan plots illustrating genetic overlap of the multivariate functional brain phenotypes with vitamin D (N = 79,366).

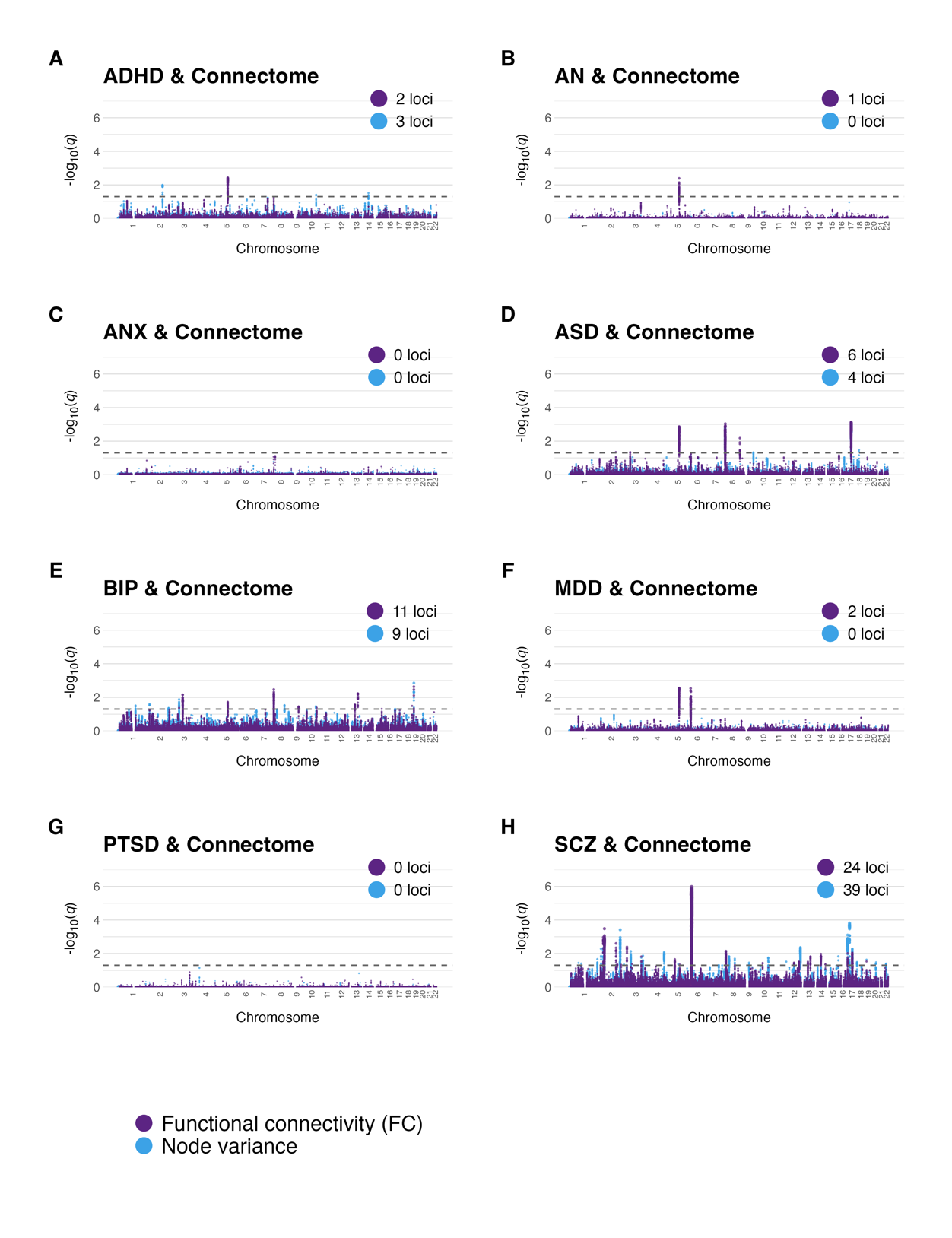

**Suppl. Fig 6. Conjunctional FDR results from the ROI-based analysis.** Manhattan plots illustrating genetic overlap between each of the psychiatric disorders (A-H) and the ROI-based imaging features.

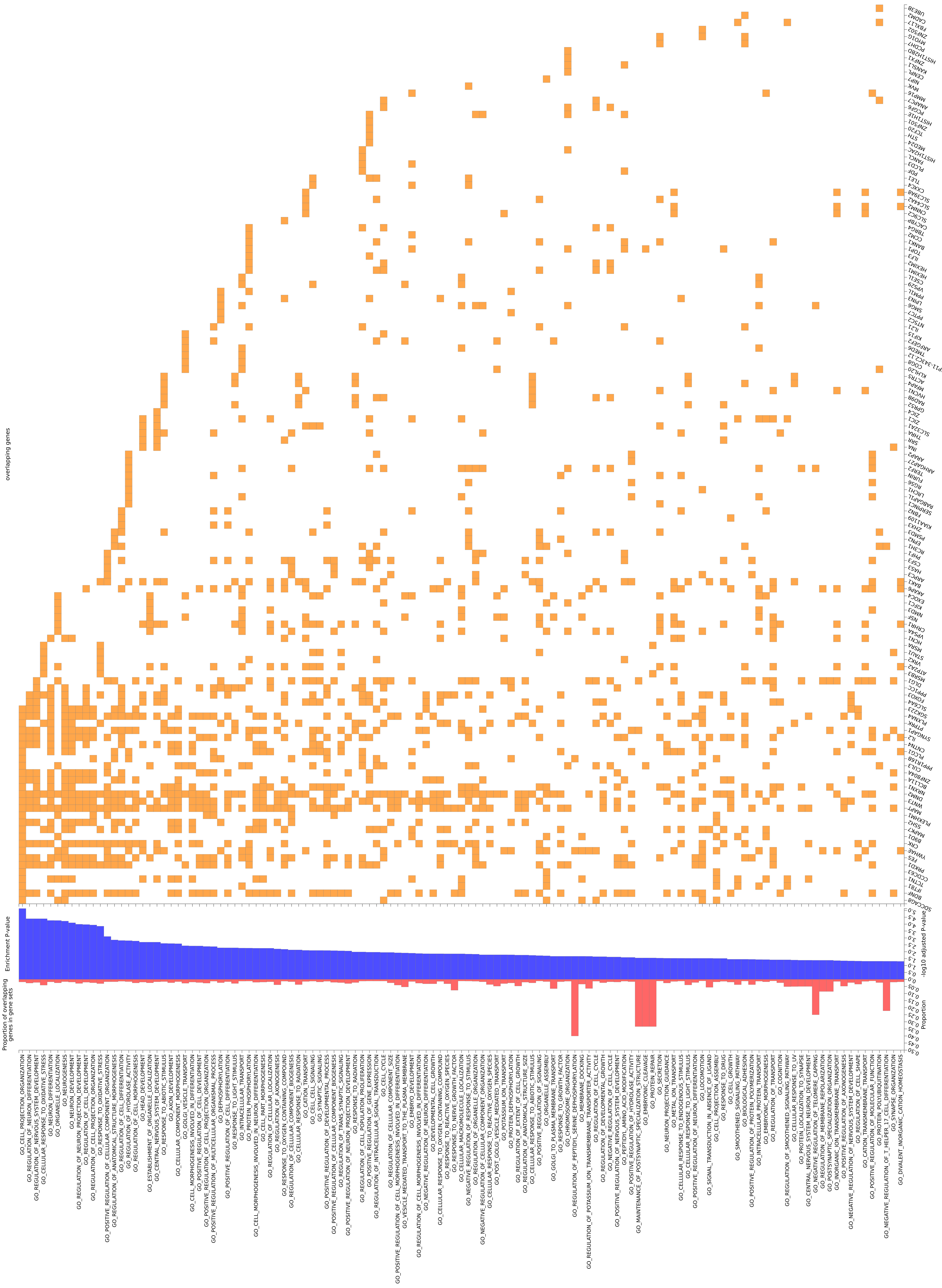

**Suppl. Fig 7. Significant enrichment of biological processes (GO).** Using FUMA gene2func, we identified 125 significant processes, many of which are relevant to neural system development and functioning, including synaptic signaling.

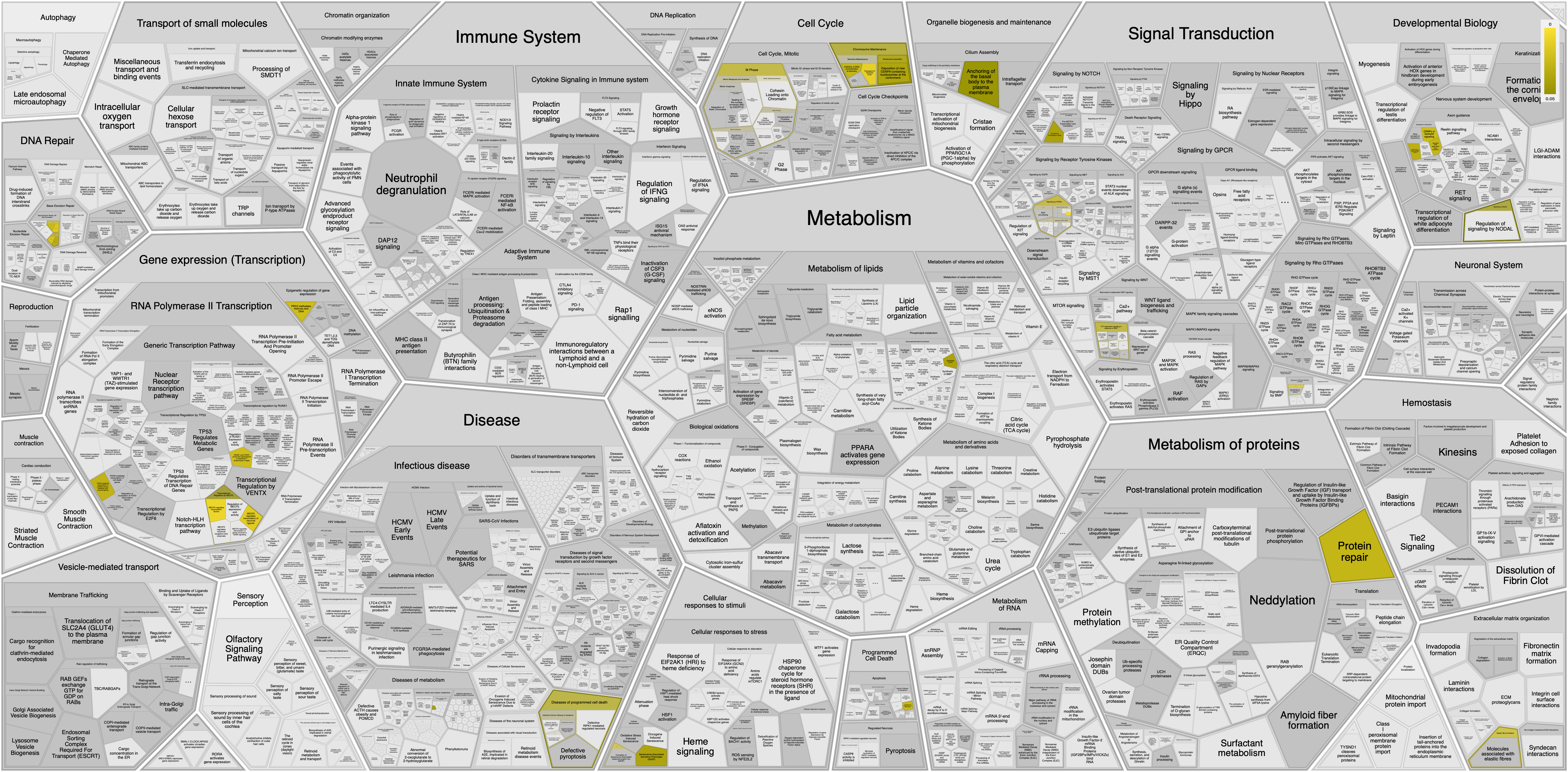

**Suppl. Fig 8.** **Voronoi visualization of mapped genes and their pathways.** Visualization of the pathways involving the mapped genes of the conjunctional FDR summary statistics from all comparisons. Figure generated by reactome.org.

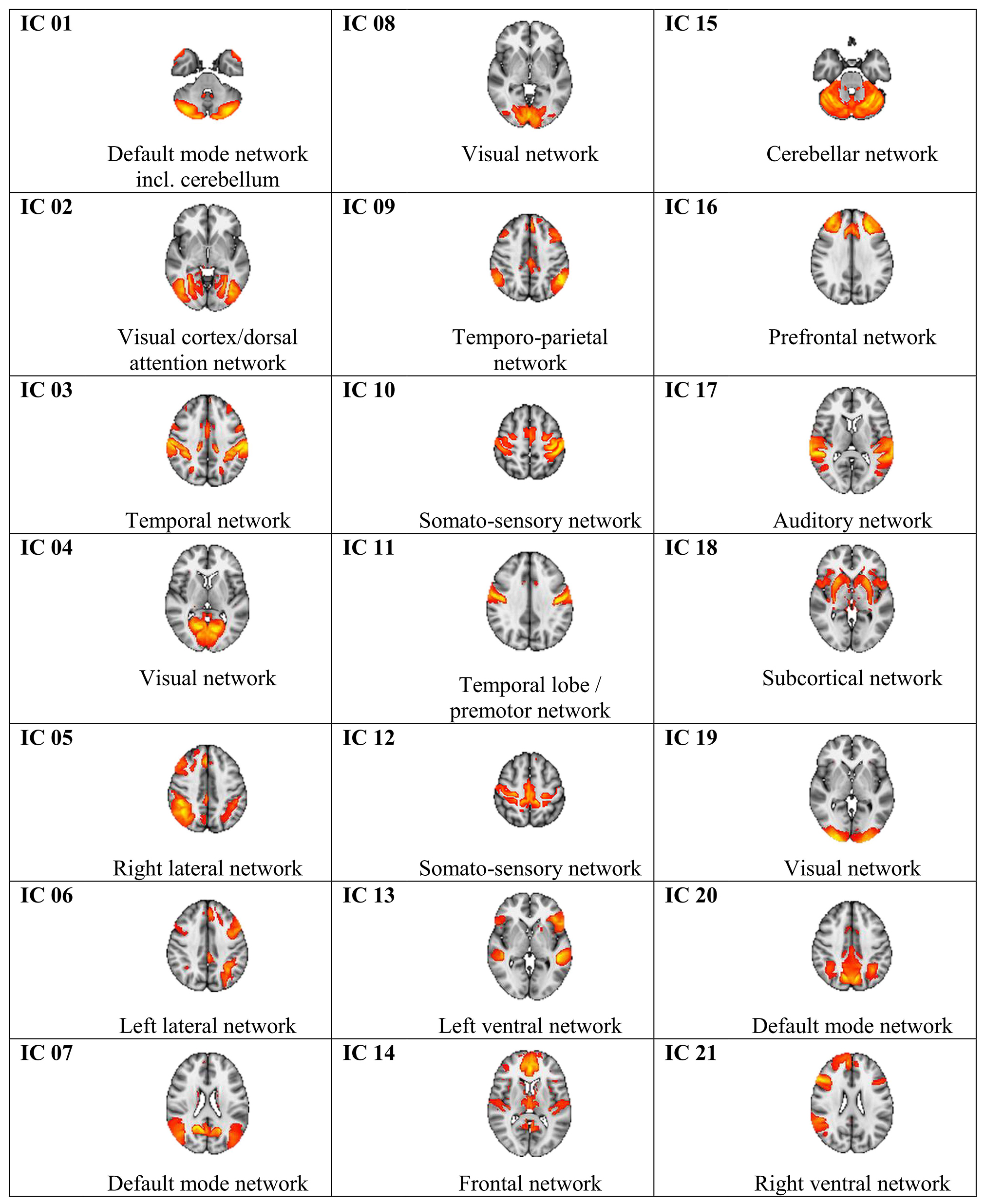

**Suppl. Fig 9.** **Maps corresponding to the 21 independent components.** For each ICA-derived brain network node, the figure shows a map alongside a descriptive label what these networks reflect. Labels are meant for an overview and do not necessarily cover all aspects of the networks.
