## Supplementary Tables for "Genetic overlap between multivariate measures of human functional brain connectivity and psychiatric disorders"

**Suppl. Table 1.** **Cohort Overview.** GWASs of psychiatric disorders included in this work. For the anorexia heritability estimate, the effective sample size was estimated using the formula 4 / (1/N_case_ + 1/N_control_).

| **Pheno-**  **type** | **Consortium** | **Sample** | **Citation** | **N_case_** | **N_control_** | **Number of loci** | **LambdaGC** | **h2**  **(se)** |
| --- | --- | --- | --- | --- | --- | --- | --- | --- |
| SCZ | PGC | Meta analysis of CLOZUK and PGC samples | Pardiñas et al., 2018 | 40,675 | 64,643 | 135 | 1.6831 | 0.41 (0.01) |
| BIP | PGC | European individuals | Mullins et al., 2021 | 41,917 | 371,549 | 44 | 1.4316 | 0.0853 (<0.01) |
| MDD | PGC | European individuals | Wray et al., 2018 | 69,576 | 161,613 | 2 | 1.1973 | 0.0717 (<0.01) |
| ADHD | PGC | European individuals | Demontis et al., 2019 | 19,099 | 34,194 | 12 | 1.2531 | 0.2354 (0.02) |
| ASD | PGC, iPSYCH | Meta analysis of European individuals | Grove et al., 2019 | 18,381 | 27,969 | 2 | 1.1715 | 0.1941 (0.02) |
| PTSD | PGC | European individuals | Duncan et al., 2018 | 2,424 | 7,113 | 0 | 1.0165 | 0.101 (0.04) |
| ANX | ANGST | Meta analysis of European individuals | Otowa et al., 2016 | 7,016 | 14,745 | 1 | 1.0345 | 0.0763 (0.03) |
| AN | PGC | European individuals | Duncan et al., 2017 | 3,495 | 10,982 | 1 | 1,0772 | 0.3281 (0.05) |
| VITD | SUNLIGHT | European individuals | Jiang et al., 2018 | 79,366 | | 6 | 1.0926 | 0.0754 (0.02) |

**Suppl. Table 2. Loci discovered using MOSTest in the main (ICA-based) analysis.**

| **Feature** | **Locus number** | **Lead SNP** | **Location** | **A1** | **A2** | **Min. BP locus** | **Max. BP locus** | **P** | **P (repli-cation)** |
| --- | --- | --- | --- | --- | --- | --- | --- | --- | --- |
| FC | 1 | rs1452628 | 1:215139887 | T | A | 215,134,041 | 215,153,067 | 3.02e-10 | 1.82e-04 |
| FC | 2 | rs2678871 | 2:58153602 | G | A | 57,942,987 | 58,484,172 | 2.22e-08 | 3.40e-02 |
| FC | 3 | rs2863957 | 2:114089551 | A | C | 114,065,390 | 114,110,568 | 3.02e-37 | 3.78e-13 |
| FC | 4 | rs11921985 | 3:89467357 | G | A | 89,273,019 | 90,012,103 | 3.55e-23 | 4.87e-08 |
| FC | 5 | rs1392261 | 3:115144685 | G | T | 114,925,815 | 115,219,766 | 3.81e-09 | 4.53e-02 |
| FC | 6 | rs2279829 | 3:147106319 | T | C | 147,020,326 | 147,224,629 | 1.50e-12 | 1.09e-05 |
| FC | 7 | rs3849198 | 6:97036539 | A | C | 96,841,762 | 97,067,047 | 1.26e-08 | 2.46e-02 |
| FC | 8 | rs2644297 | 7:2858906 | C | A | 2,752,152 | 2,912,928 | 2.21e-09 | 3.88e-03 |
| FC | 9 | rs57866767 | 10:96023077 | C | T | 95,988,042 | 96,127,448 | 3.01e-18 | 8.56e-10 |
| FC | 10 | rs11819412 | 10:134297803 | G | C | 134,295,388 | 134,335,986 | 1.36e-11 | 9.43e-03 |
| FC | 11 | rs977671 | 11:80045121 | A | G | 80,044,823 | 80,074,551 | 1.31e-09 | 5.96e-03 |
| FC | 12 | rs653178 | 12:112007756 | C | T | 111,826,477 | 112,906,415 | 3.63e-08 | 1.26e-01 |
| FC | 13 | rs62056163 | 16:87255310 | G | A | 87,222,153 | 87,262,642 | 3.52e-10 | 6.79e-05 |
| FC | 14 | rs62055701 | 17:43758787 | A | G | 43,460,181 | 44,865,498 | 2.50e-23 | 8.69e-05 |
| FC | 15 | rs429358 | 19:45411941 | C | T | 45,386,467 | 45,428,234 | 1.46e-30 | 2.17e-09 |
| Node variance | 1 | rs61784835 | 1:47974123 | T | C | 47,974,123 | 47,980,916 | 1.27e-08 | 4.56e-02 |
| Node variance | 2 | rs6737318 | 2:114083120 | G | A | 114,065,390 | 114,123,988 | 2.99e-49 | 1.26e-11 |
| Node variance | 3 | rs56023709 | 16:87229344 | A | C | 87,221,646 | 87,258,088 | 6.12e-10 | 1.79e-03 |
| Node variance | 4 | rs71375338 | 17:44332793 | A | G | 43,572,419 | 44,865,603 | 4.74e-10 | 3.67e-03 |
| Node variance | 5 | rs429358 | 19:45411941 | C | T | 45,386,467 | 45,428,234 | 1.87e-28 | 7.74e-08 |

**Suppl. Table 3. Loci discovered using MOSTest in the region-of-interest (ROI)-based analysis.**

| **Feature** | **Locus number** | **Lead SNP** | **Location** | **A1** | **A2** | **Min. BP locus** | **Max. BP locus** | **P** |
| --- | --- | --- | --- | --- | --- | --- | --- | --- |
| FC | 1 | rs6658111 | 1:47980916 | G | T | 47,974,123 | 47,980,916 | 1.99e-08 |
| FC | 2 | rs7601767 | 2:48277490 | A | G | 48,178,775 | 48,324,044 | 1.67e-12 |
| FC | 3 | rs6709720 | 2:57967563 | A | G | 57,942,987 | 58,300,783 | 2.17e-08 |
| FC | 4 | rs2863957 | 2:114089551 | A | C | 114,065,390 | 114,110,568 | 7.99e-35 |
| FC | 5 | rs10930040 | 2:162894766 | G | A | 162,797,621 | 162,908,664 | 1.65e-08 |
| FC | 6 | rs13317478 | 3:17464862 | C | T | 17,304,712 | 17,747,914 | 1.02e-08 |
| FC | 7 | rs9814516 | 3:85407980 | T | G | 85,397,049 | 85,791,383 | 1.14e-11 |
| FC | 8 | rs35124509 | 3:89521693 | C | T | 89,041,916 | 90,502,283 | 1.32e-59 |
| FC | 9 | rs2279829 | 3:147106319 | T | C | 147,090,583 | 147,268,599 | 8.68e-17 |
| FC | 10 | rs114468556 | 5:92870246 | A | T | 92,683,607 | 93,537,741 | 5.06e-12 |
| FC | 11 | rs3094222 | 6:31081434 | G | A | 29,752,808 | 32,205,942 | 9.48e-10 |
| FC | 12 | rs1182175 | 7:2875026 | G | A | 2,769,921 | 2,901,084 | 4.67e-08 |
| FC | 13 | rs615632 | 8:9796321 | C | T | 8,543,324 | 10,193,772 | 4.02e-09 |
| FC | 14 | rs3891783 | 10:96015793 | G | C | 96,009,182 | 96,069,405 | 2.28e-08 |
| FC | 15 | rs2559509 | 10:126551900 | C | A | 126,384,610 | 126,556,015 | 3.85e-15 |
| FC | 16 | rs10892823 | 11:122128720 | G | T | 122,089,589 | 122,198,078 | 3.28e-09 |
| FC | 17 | rs160459 | 14:59074136 | C | A | 59,064,730 | 59,092,581 | 3.59e-08 |
| FC | 18 | rs2164950 | 14:59627631 | A | G | 59,588,323 | 59,669,948 | 1.61e-12 |
| FC | 19 | rs75022332 | 17:43810873 | T | A | 43,463,493 | 44,809,001 | 2.73e-12 |
| FC | 20 | rs73006822 | 19:13109955 | T | C | 13,089,890 | 13,165,884 | 2.71e-09 |
| FC | 21 | rs2043294 | 19:32207820 | T | A | 32,200,518 | 32,208,909 | 6.02e-12 |
| FC | 22 | rs429358 | 19:45411941 | C | T | 45,392,254 | 45,428,234 | 1.24e-16 |
| Node variance | 1 | rs2066981 | 1:155172379 | G | A | 155,123,837 | 155,197,462 | 4.55e-08 |
| Node variance | 2 | rs62158169 | 2:114081827 | T | C | 114,065,390 | 114,123,988 | 4.44e-37 |
| Node variance | 3 | rs35124509 | 3:89521693 | C | T | 89,451,721 | 89,751,353 | 5.96e-09 |
| Node variance | 4 | rs2517718 | 6:29916391 | C | A | 29,804,165 | 29,924,389 | 1.71e-08 |
| Node variance | 5 | rs7748505 | 6:45316249 | C | A | 44,673,787 | 45,404,230 | 4.81e-16 |
| Node variance | 6 | rs72932805 | 6:96902933 | G | T | 96,767,685 | 97,067,047 | 1.74e-14 |
| Node variance | 7 | rs1636264 | 7:2864586 | T | G | 2,752,152 | 2,912,928 | 2.60e-12 |
| Node variance | 8 | rs55766546 | 7:14741152 | G | C | 14,661,337 | 14,745,441 | 2.80e-13 |
| Node variance | 9 | rs16896921 | 8:99369325 | T | C | 99,364,152 | 99,383,449 | 3.59e-14 |
| Node variance | 10 | rs4335155 | 8:120013469 | A | C | 119,904,557 | 120,063,542 | 6.73e-11 |
| Node variance | 11 | rs7042786 | 9:71431174 | A | T | 71,397,747 | 71,475,381 | 3.15e-12 |
| Node variance | 12 | rs1243186 | 10:21902760 | A | G | 21,768,560 | 22,288,132 | 6.81e-11 |
| Node variance | 13 | rs57866767 | 10:96023077 | C | T | 95,971,321 | 96,133,084 | 7.30e-36 |
| Node variance | 14 | rs7080018 | 10:134301505 | G | A | 134,269,304 | 134,335,986 | 1.13e-24 |
| Node variance | 15 | rs753002 | 11:10739243 | G | A | 10,704,733 | 10,748,069 | 1.22e-11 |
| Node variance | 16 | rs112866565 | 11:26232313 | G | T | 26,164,646 | 26,278,200 | 2.98e-08 |
| Node variance | 17 | rs7479309 | 11:27311814 | A | G | 27,265,644 | 27,338,712 | 1.88e-10 |
| Node variance | 18 | rs12807936 | 11:69998470 | T | C | 69,952,562 | 70,007,484 | 8.19e-09 |
| Node variance | 19 | rs6590919 | 11:101646567 | T | C | 101,646,567 | 101,816,307 | 4.70e-09 |
| Node variance | 20 | rs7134473 | 12:28340578 | G | A | 28,262,880 | 28,698,528 | 1.22e-11 |
| Node variance | 21 | rs10784446 | 12:65764979 | A | G | 65,762,058 | 65,883,743 | 2.44e-08 |
| Node variance | 22 | rs9899649 | 17:19253613 | C | T | 19,138,174 | 19,299,144 | 9.84e-12 |
| Node variance | 23 | rs429358 | 19:45411941 | C | T | 45,392,254 | 45,428,234 | 1.46e-15 |
| Node variance | 24 | rs41298840 | 22:19753449 | G | A | 19,750,773 | 19,765,181 | 1.54e-08 |

**Suppl. Table 4. Number of loci significantly overlapping between the psychiatric disorders and functional connectivity / node variance.** The main analysis was performed using the standard 5% threshold for FDR correction. A sensitivity analysis with a more stringent 1% threshold confirmed largest overlap for SCZ with FC among the traits.

| **fMRI feature** | **Diagnosis** | **Number of loci**  **(FDR level 5%)** | **Number of loci**  **(FDR level 1%)** |
| --- | --- | --- | --- |
| connectivity | ADHD | 6 | 0 |
| node variance | ADHD | 1 | 0 |
| connectivity | AN | 1 | 1 |
| node variance | AN | 0 | 0s |
| connectivity | ANX | 0 | 0 |
| node variance | ANX | 1 | 0 |
| connectivity | ASD | 4 | 2 |
| node variance | ASD | 3 | 1 |
| connectivity | BIP | 9 | 3 |
| node variance | BIP | 2 | 0 |
| connectivity | MDD | 1 | 0 |
| node variance | MDD | 0 | 0 |
| connectivity | PTSD | 0 | 0 |
| node variance | PTSD | 0 | 0 |
| connectivity | SCZ | 43 | 10 |
| node variance | SCZ | 22 | 2 |

**Suppl. Table 5. Mapped genes from FUMA.** This table shows the mapped genes and individual lead SNPs as provided by FUMA. A total of 180 unique genes were mapped for 5 of the 8 disorders analyzed in this manuscript.

| **Diagnosis** | **Feature** | **No** | **Gene** | **Chr** | **Pmin** | **Ind. Sig. SNP** |
| --- | --- | --- | --- | --- | --- | --- |
| ADHD | edge | 1 | FBXL17 | 5 | 4.799e-07 | rs289230 |
| ADHD | edge | 2 | AKAP6 | 14 | 4.789e-07 | rs2383378 |
| ANX | node | 1 | SLC9C2 | 1 | 5.882e-06 | rs61826888 |
| ANX | node | 2 | ANKRD45 | 1 | 4.998e-06 | rs61826888 |
| ANX | node | 3 | KLHL20 | 1 | 7.006e-06 | rs61826888 |
| ANX | node | 4 | CENPL | 1 | 9.997e-06 | rs61826888 |
| ANX | node | 5 | DARS2 | 1 | 9.997e-06 | rs61826888 |
| ANX | node | 6 | ZBTB37 | 1 | 9.154e-06 | rs61826888 |
| ANX | node | 7 | SERPINC1 | 1 | 8.826e-06 | rs61826888 |
| ANX | node | 8 | RC3H1 | 1 | 3.932e-06 | rs61826888 |
| ANX | node | 9 | RABGAP1L | 1 | 4.655e-07 | rs61826888 |
| ANX | node | 10 | GPR52 | 1 | 9.778e-06 | rs61826888 |
| ANX | node | 11 | CACYBP | 1 | 9.662e-06 | rs61826888 |
| ANX | node | 12 | MRPS14 | 1 | 9.662e-06 | rs61826888 |
| ASD | edge | 1 | CXXC4 | 4 | 2.342e-07 | rs4475153 |
| ASD | edge | 2 | XKR6 | 8 | 1.029e-08 | rs11250093 |
| ASD | edge | 3 | PLEKHM1 | 17 | 2.286e-08 | rs71375338 |
| ASD | edge | 4 | CRHR1 | 17 | 1.258e-08 | rs71375338 |
| ASD | edge | 5 | SPPL2C | 17 | 1.030e-08 | rs71375338 |
| ASD | edge | 6 | MAPT | 17 | 1.043e-08 | rs71375338 |
| ASD | edge | 7 | STH | 17 | 1.594e-08 | rs71375338 |
| ASD | edge | 8 | KANSL1 | 17 | 8.417e-09 | rs71375338 |
| ASD | edge | 9 | ARL17B | 17 | 8.670e-09 | rs71375338 |
| ASD | edge | 10 | LRRC37A | 17 | 1.861e-08 | rs71375338 |
| ASD | edge | 11 | LRRC37A2 | 17 |  | rs71375338 |
| ASD | edge | 12 | ARL17A | 17 |  | rs71375338 |
| ASD | edge | 13 | NSF | 17 | 7.823e-08 | rs71375338 |
| ASD | node | 1 | FBN2 | 5 | 4.813e-07 | rs11745497 |
| ASD | node | 2 | PLEKHM1 | 17 | 2.431e-08 | rs71375338 |
| ASD | node | 3 | CRHR1 | 17 | 1.302e-08 | rs71375338 |
| ASD | Mnode | 4 | SPPL2C | 17 | 1.056e-08 | rs71375338 |
| ASD | node | 5 | MAPT | 17 | 1.070e-08 | rs71375338 |
| ASD | node | 6 | STH | 17 | 1.669e-08 | rs71375338 |
| ASD | node | 7 | KANSL1 | 17 | 8.551e-09 | rs71375338 |
| ASD | node | 8 | ARL17B | 17 | 8.819e-09 | rs71375338 |
| ASD | node | 9 | LRRC37A | 17 | 1.962e-08 | rs71375338 |
| ASD | node | 10 | LRRC37A2 | 17 |  | rs71375338 |
| ASD | node | 11 | ARL17A | 17 |  | rs71375338 |
| ASD | node | 12 | NSF | 17 | 8.453e-08 | rs71375338 |
| BIP | edge | 1 | ZNF502 | 3 | 4.662e-07 | rs3804580 |
| BIP | edge | 2 | ZNF501 | 3 | 4.662e-07 | rs3804580 |
| BIP | edge | 3 | KIAA1143 | 3 | 5.647e-07 | rs3804580 |
| BIP | edge | 4 | KIF15 | 3 | 3.969e-07 | rs3804580 |
| BIP | edge | 5 | TMEM42 | 3 | 4.969e-07 | rs3804580 |
| BIP | edge | 6 | TGM4 | 3 | 4.969e-07 | rs3804580 |
| BIP | edge | 7 | CADM2 | 3 | 7.160e-08 | rs62250713 |
| BIP | edge | 8 | MYO1G | 7 | 1.109e-06 | rs2304693 |
| BIP | edge | 9 | CCM2 | 7 | 1.553e-06 | rs2304693 |
| BIP | edge | 10 | NACAD | 7 | 1.511e-06 | rs2304693 |
| BIP | edge | 11 | TBRG4 | 7 | 4.614e-07 | rs2304693 |
| BIP | edge | 12 | MSRA | 8 | 1.378e-06 | rs615632 |
| BIP | edge | 13 | HS6ST3 | 13 | 4.877e-07 | rs7996549 |
| BIP | edge | 14 | FURIN | 15 | 4.977e-08 | rs4702 |
| BIP | edge | 15 | FES | 15 | 4.977e-08 | rs4702 |
| BIP | edge | 16 | GSDMA | 17 | 3.601e-06 | rs10445310 |
| BIP | edge | 17 | PSMD3 | 17 | 3.580e-06 | rs10445310 |
| BIP | edge | 18 | CSF3 | 17 | 1.418e-06 | rs10445310 |
| BIP | edge | 19 | MED24 | 17 | 2.400e-07 | rs10445310 |
| BIP | edge | 20 | THRA | 17 | 2.400e-07 | rs10445310 |
| BIP | edge | 21 | SLC44A2 | 19 | 7.236e-08 | rs1053007 |
| BIP | edge | 22 | AC011475.1 | 19 | 7.236e-08 | rs1053007 |
| BIP | edge | 23 | ILF3 | 19 | 8.157e-08 | rs1053007 |
| BIP | edge | 24 | QTRT1 | 19 | 8.506e-08 | rs1053007 |
| BIP | edge | 25 | DNM2 | 19 | 5.386e-07 | rs1053007 |
| BIP | node | 1 | ARAP2 | 4 | 4.370e-07 | rs34680259 |
| BIP | node | 2 | PLXNA4 | 7 | 2.058e-07 | rs3734989 |
| SCZ | edge | 1 | CEP170 | 1 | 2.539e-06 | rs10926994 |
| SCZ | edge | 2 | AC092782.1 | 1 | 3.763e-06 | rs10926994 |
| SCZ | edge | 3 | SDCCAG8 | 1 | 4.217e-07 | rs10926994 |
| SCZ | edge | 4 | NRXN1 | 2 | 2.842e-07 | rs116748071 |
| SCZ | edge | 5 | VRK2 | 2 | 1.832e-10 | rs2678871 |
| SCZ | edge | 6 | FANCL | 2 | 3.192e-07 | rs2678871 |
| SCZ | edge | 7 | BCL11A | 2 | 3.255e-07 | rs13019832 |
| SCZ | edge | 8 | ZNF804A | 2 | 3.504e-07 | rs1583048 |
| SCZ | edge | 9 | CNTN4 | 3 | 2.630e-07 | rs6778940 |
| SCZ | edge | 10 | EXOG | 3 | 1.620e-07 | rs11717146 |
| SCZ | edge | 11 | RBM6 | 3 |  | rs2624847 |
| SCZ | edge | 12 | ZIC4 | 3 | 9.328e-08 | rs2279829 |
| SCZ | edge | 13 | ZIC1 | 3 | 9.328e-08 | rs2279829 |
| SCZ | edge | 14 | PPM1L | 3 | 2.414e-06 | rs465985 |
| SCZ | edge | 15 | B3GALNT1 | 3 | 1.817e-07 | rs465985 |
| SCZ | edge | 16 | NMD3 | 3 | 1.657e-07 | rs465985 |
| SCZ | edge | 17 | SPTSSB | 3 | 5.916e-08 | rs465985 |
| SCZ | edge | 18 | PCDH7 | 4 | 2.366e-07 | rs11721555 |
| SCZ | edge | 19 | BANK1 | 4 | 2.678e-06 | rs13107325 |
| SCZ | edge | 20 | SLC39A8 | 4 | 3.822e-09 | rs13107325 |
| SCZ | edge | 21 | CXXC4 | 4 | 2.030e-07 | rs13111612 |
| SCZ | edge | 22 | KIAA1109 | 4 | 3.546e-07 | rs77087420 |
| SCZ | edge | 23 | ADAD1 | 4 |  | rs77087420 |
| SCZ | edge | 24 | IL2 | 4 |  | rs77087420 |
| SCZ | edge | 25 | IL21 | 4 | 2.104e-06 | rs77087420 |
| SCZ | edge | 26 | HCN1 | 5 | 5.198e-06 | rs13177136 |
| SCZ | edge | 27 | HIST1H2AC | 6 | 2.776e-07 | rs9379829 |
| SCZ | edge | 28 | HIST1H1E | 6 | 1.379e-07 | rs9379829 |
| SCZ | edge | 29 | HIST1H2BD | 6 | 5.002e-08 | rs9379829 |
| SCZ | edge | 30 | SCAND3 | 6 | 2.784e-08 | rs9357070 |
| SCZ | edge | 31 | KIFC1 | 6 | 3.768e-07 | rs9278045 |
| SCZ | edge | 32 | PHF1 | 6 | 3.768e-07 | rs9278045 |
| SCZ | edge | 33 | CUTA | 6 | 3.903e-07 | rs9278045 |
| SCZ | edge | 34 | SYNGAP1 | 6 | 3.903e-07 | rs9278045 |
| SCZ | edge | 35 | ZBTB9 | 6 | 7.751e-07 | rs9278045 |
| SCZ | edge | 36 | BAK1 | 6 | 5.453e-06 | rs9278045 |
| SCZ | edge | 37 | PTPRK | 6 | 3.183e-07 | rs7742212 |
| SCZ | edge | 38 | EXOC4 | 7 | 3.365e-07 | rs12539120 |
| SCZ | edge | 39 | SGK223 | 8 | 4.342e-06 | rs2945232 |
| SCZ | edge | 40 | TLE1 | 9 | 4.534e-07 | rs9410573 |
| SCZ | edge | 41 | WBP1L | 10 | 2.668e-06 | rs113278154 |
| SCZ | edge | 42 | CYP17A1 | 10 | 1.460e-06 | rs113278154 |
| SCZ | edge | 43 | C10orf32 | 10 | 1.113e-06 | rs113278154 |
| SCZ | edge | 44 | C10orf32-ASMT | 10 | 5.641e-07 | rs113278154 |
| SCZ | edge | 45 | AS3MT | 10 | 5.641e-07 | rs113278154 |
| SCZ | edge | 46 | CNNM2 | 10 | 3.147e-07 | rs113278154 |
| SCZ | edge | 47 | NT5C2 | 10 | 2.249e-07 | rs113278154 |
| SCZ | edge | 48 | INA | 10 | 2.071e-06 | rs113278154 |
| SCZ | edge | 49 | PCGF6 | 10 | 2.071e-06 | rs113278154 |
| SCZ | edge | 50 | BDNF | 11 | 9.643e-08 | rs11030104 |
| SCZ | edge | 51 | MSRB3 | 12 | 2.312e-07 | rs10878264 |
| SCZ | edge | 52 | MYO1H | 12 | 3.461e-07 | rs7953142 |
| SCZ | edge | 53 | KCTD10 | 12 | 5.996e-07 | rs7953142 |
| SCZ | edge | 54 | UBE3B | 12 |  | rs7953142 |
| SCZ | edge | 55 | MMAB | 12 |  | rs7953142 |
| SCZ | edge | 56 | MVK | 12 | 1.869e-06 | rs7953142 |
| SCZ | edge | 57 | C12orf76 | 12 | 2.905e-06 | rs60027620 |
| SCZ | edge | 58 | IFT81 | 12 | 1.219e-06 | rs60027620 |
| SCZ | edge | 59 | ATP2A2 | 12 | 6.671e-07 | rs60027620 |
| SCZ | edge | 60 | ANAPC7 | 12 | 8.131e-07 | rs60027620 |
| SCZ | edge | 61 | ARPC3 | 12 | 4.419e-07 | rs60027620 |
| SCZ | edge | 62 | GPN3 | 12 | 6.583e-07 | rs60027620 |
| SCZ | edge | 63 | FAM216A | 12 | 6.369e-07 | rs60027620 |
| SCZ | edge | 64 | VPS29 | 12 | 6.369e-07 | rs60027620 |
| SCZ | edge | 65 | RAD9B | 12 | 6.606e-07 | rs60027620 |
| SCZ | edge | 66 | PPTC7 | 12 | 5.749e-07 | rs60027620 |
| SCZ | edge | 67 | TCTN1 | 12 | 4.734e-07 | rs60027620 |
| SCZ | edge | 68 | HVCN1 | 12 | 3.450e-07 | rs60027620 |
| SCZ | edge | 69 | PPP1CC | 12 | 4.650e-07 | rs60027620 |
| SCZ | edge | 70 | CCDC63 | 12 | 6.840e-07 | rs60027620 |
| SCZ | edge | 71 | LRCH1 | 13 | 3.764e-07 | rs1324007 |
| SCZ | edge | 72 | PRKD1 | 14 | 4.940e-07 | rs45438099 |
| SCZ | edge | 73 | AKAP6 | 14 | 4.449e-07 | rs2300861 |
| SCZ | edge | 74 | RGS6 | 14 | 1.568e-07 | rs71427144 |
| SCZ | edge | 75 | AC005477.1 | 14 | 1.568e-07 | rs71427144 |
| SCZ | edge | 76 | FURIN | 15 | 2.361e-08 | rs4702 |
| SCZ | edge | 77 | FES | 15 | 2.361e-08 | rs4702 |
| SCZ | edge | 78 | HAS3 | 16 | 2.765e-06 | rs12925547 |
| SCZ | edge | 79 | CHTF8 | 16 | 3.731e-06 | rs12925547 |
| SCZ | edge | 80 | CHTF8 | 16 | 5.446e-06 | rs12925547 |
| SCZ | edge | 81 | CIRH1A | 16 | 3.970e-07 | rs12925547 |
| SCZ | edge | 82 | SNTB2 | 16 | 3.970e-07 | rs12925547 |
| SCZ | edge | 83 | RP11-343C2.11 | 16 | 6.140e-07 | rs12925547 |
| SCZ | edge | 84 | VPS4A | 16 | 6.522e-07 | rs12925547 |
| SCZ | edge | 85 | COG8 | 16 | 7.113e-07 | rs12925547 |
| SCZ | edge | 86 | PDF | 16 | 7.553e-07 | rs12925547 |
| SCZ | edge | 87 | RP11-343C2.12 | 16 | 7.553e-07 | rs12925547 |
| SCZ | edge | 88 | RP11-343C2.9 | 16 | 1.155e-06 | rs12925547 |
| SCZ | edge | 89 | RP11-343C2.7 | 16 | 1.155e-06 | rs12925547 |
| SCZ | edge | 90 | NIP7 | 16 | 1.155e-06 | rs12925547 |
| SCZ | edge | 91 | TMED6 | 16 | 1.155e-06 | rs12925547 |
| SCZ | edge | 92 | TERF2 | 16 | 2.040e-06 | rs12925547 |
| SCZ | edge | 93 | YWHAE | 17 | 4.861e-07 | rs58143069 |
| SCZ | edge | 94 | CRK | 17 | 6.685e-07 | rs58143069 |
| SCZ | edge | 95 | EPN2 | 17 | 1.436e-07 | rs1964703 |
| SCZ | edge | 96 | B9D1 | 17 | 1.106e-07 | rs1964703 |
| SCZ | edge | 97 | MAPK7 | 17 | 2.859e-06 | rs1964703 |
| SCZ | edge | 98 | MFAP4 | 17 | 2.859e-06 | rs1964703 |
| SCZ | edge | 99 | PLCD3 | 17 | 2.118e-07 | rs60289499 |
| SCZ | edge | 100 | ACBD4 | 17 | 2.118e-07 | rs60289499 |
| SCZ | edge | 101 | HEXIM1 | 17 | 2.118e-07 | rs60289499 |
| SCZ | edge | 102 | HEXIM2 | 17 | 3.063e-07 | rs60289499 |
| SCZ | edge | 103 | ARHGAP27 | 17 | 2.511e-06 | rs2532392 |
| SCZ | edge | 104 | PLEKHM1 | 17 | 4.939e-07 | rs2532392 |
| SCZ | edge | 105 | CRHR1 | 17 | 1.435e-07 | rs2532392 |
| SCZ | edge | 106 | SPPL2C | 17 | 1.447e-07 | rs2532392 |
| SCZ | edge | 107 | MAPT | 17 | 6.671e-08 | rs2532392 |
| SCZ | edge | 108 | STH | 17 | 1.135e-07 | rs2532392 |
| SCZ | edge | 109 | KANSL1 | 17 | 5.154e-08 | rs2532392 |
| SCZ | edge | 110 | ARL17B | 17 | 6.924e-08 | rs2532392 |
| SCZ | edge | 111 | LRRC37A | 17 | 1.777e-07 | rs2532392 |
| SCZ | edge | 112 | LRRC37A2 | 17 |  | rs2532392 |
| SCZ | edge | 113 | ARL17A | 17 |  | rs2532392 |
| SCZ | edge | 114 | NSF | 17 | 1.597e-07 | rs2532392 |
| SCZ | edge | 115 | WNT3 | 17 | 1.050e-07 | rs2532392 |
| SCZ | edge | 116 | ZNF407 | 18 | 4.177e-07 | rs9989571 |
| SCZ | edge | 117 | SLC32A1 | 20 | 3.099e-06 | rs12481241 |
| SCZ | edge | 118 | ACTR5 | 20 |  | rs12481241 |
| SCZ | edge | 119 | PPP1R16B | 20 | 3.813e-07 | rs12481241 |
| SCZ | node | 1 | ZBTB37 | 1 | 2.716e-07 | rs4592301 |
| SCZ | node | 2 | SERPINC1 | 1 | 2.716e-07 | rs4592301 |
| SCZ | node | 3 | RC3H1 | 1 | 1.804e-07 | rs4592301 |
| SCZ | node | 4 | RABGAP1L | 1 | 9.459e-08 | rs4592301 |
| SCZ | node | 5 | VRK2 | 2 | 4.398e-07 | rs2678871;rs34556232 |
| SCZ | node | 6 | FANCL | 2 | 3.362e-07 | rs34556232;rs2678871 |
| SCZ | node | 7 | CUL3 | 2 | 4.578e-07 | rs11676311 |
| SCZ | node | 8 | ZIC4 | 3 | 1.011e-07 | rs2279829 |
| SCZ | node | 9 | ZIC1 | 3 | 1.011e-07 | rs2279829 |
| SCZ | node | 10 | DLG1 | 3 | 4.427e-07 | rs10489880 |
| SCZ | node | 11 | BANK1 | 4 | 2.247e-07 | rs13109404 |
| SCZ | node | 12 | FOXO3 | 6 | 3.746e-07 | rs4946935 |
| SCZ | node | 13 | MMP16 | 8 | 1.490e-06 | rs3110417 |
| SCZ | node | 14 | WBP1L | 10 | 1.373e-06 | rs7085104 |
| SCZ | node | 15 | CYP17A1 | 10 | 7.716e-07 | rs7085104 |
| SCZ | node | 16 | C10orf32 | 10 | 2.914e-07 | rs7085104 |
| SCZ | node | 17 | C10orf32-ASMT | 10 | 2.914e-07 | rs7085104 |
| SCZ | node | 18 | AS3MT | 10 | 2.914e-07 | rs7085104 |
| SCZ | node | 19 | CNNM2 | 10 | 3.709e-07 | rs7085104 |
| SCZ | node | 20 | NT5C2 | 10 | 5.885e-07 | rs7085104 |
| SCZ | node | 21 | RBFOX1 | 16 | 2.115e-07 | rs810609 |
| SCZ | node | 22 | SMG6 | 17 | 2.502e-07 | rs9303272 |
| SCZ | node | 23 | SRR | 17 | 6.292e-07 | rs9303272 |
| SCZ | node | 24 | SSH2 | 17 | 3.365e-06 | rs4514736 |
| SCZ | node | 25 | EFCAB5 | 17 | 2.273e-06 | rs4514736 |
| SCZ | node | 26 | NSRP1 | 17 | 1.752e-06 | rs4514736 |
| SCZ | node | 27 | SLC6A4 | 17 | 3.311e-07 | rs4514736 |
| SCZ | node | 28 | ARHGAP27 | 17 | 2.759e-06 | rs2532392 |
| SCZ | node | 29 | PLEKHM1 | 17 | 5.466e-07 | rs2532392 |
| SCZ | node | 30 | CRHR1 | 17 | 1.529e-07 | rs2532392 |
| SCZ | node | 31 | SPPL2C | 17 | 1.542e-07 | rs2532392 |
| SCZ | node | 32 | MAPT | 17 | 7.427e-08 | rs2532392 |
| SCZ | node | 33 | STH | 17 | 1.218e-07 | rs2532392 |
| SCZ | node | 34 | KANSL1 | 17 | 5.902e-08 | rs2532392 |
| SCZ | node | 35 | ARL17B | 17 | 7.682e-08 | rs2532392 |
| SCZ | node | 36 | LRRC37A | 17 | 1.891e-07 | rs2532392 |
| SCZ | node | 37 | LRRC37A2 | 17 |  | rs2532392 |
| SCZ | node | 38 | ARL17A | 17 |  | rs2532392 |
| SCZ | node | 39 | NSF | 17 | 1.700e-07 | rs2532392 |
| SCZ | node | 40 | WNT3 | 17 | 1.130e-07 | rs2532392 |
| SCZ | node | 41 | TOP1 | 20 | 7.189e-06 | rs17265513 |
| SCZ | node | 42 | PLCG1 | 20 | 7.544e-06 | rs17265513 |
| SCZ | node | 43 | ZHX3 | 20 | 2.922e-07 | rs17265513 |
| SCZ | node | 44 | LPIN3 | 20 | 9.430e-06 | rs17265513 |
| SCZ | node | 45 | ARFGEF2 | 20 | 5.442e-07 | rs6067007 |
| SCZ | node | 46 | CSE1L | 20 | 6.869e-07 | rs6067007 |
| SCZ | node | 47 | STAU1 | 20 | 5.634e-07 | rs6067007 |
| SCZ | node | 48 | DDX27 | 20 | 1.275e-06 | rs6067007 |
| SCZ | node | 49 | ZNFX1 | 20 | 1.043e-06 | rs6067007 |
| SCZ | node | 50 | TCF20 | 22 | 3.935e-07 | rs5751239 |
| SCZ | node | 51 | FLJ27365 | 22 | 2.388e-07 | rs6520050 |

**Suppl. Table 6. Mapped genes from FUMA for the ROI comparisons.** This table shows the mapped genes and individual lead SNPs as provided by FUMA for the ROI-based analyses. A total of 265 unique genes were mapped for 5 of the 8 disorders analyzed in this manuscript.

| Diagnosis | Feature | No | Gene | Chr | Pmin | Ind. Sig. SNP |
| --- | --- | --- | --- | --- | --- | --- |
| ADHD | edge | 1 | ZNF131 | 5 | 4.6e-07 | rs10041458 |
| ADHD | edge | 2 | NIM1K | 5 | 4.6e-07 | rs10041458 |
| ADHD | node | 1 | SORCS3 | 10 | 3.927e-07 | rs1027190 |
| ADHD | node | 2 | C14orf39 | 14 | 3.068e-07 | rs1272131 |
| ADHD | node | 3 | SIX6 | 14 | 4.147e-07 | rs1272131 |
| ADHD | node | 4 | SIX1 | 14 | 4.855e-07 | rs1272131 |
| ASD | edge | 1 | SLC4A10 | 2 | 1.657e-06 | rs2300755 |
| ASD | edge | 2 | DPP4 | 2 | 4.478e-07 | rs2300755 |
| ASD | edge | 3 | PRKAR2A | 3 | 1.433e-06 | rs73077200 |
| ASD | edge | 4 | SLC25A20 | 3 | 1.433e-06 | rs73077200 |
| ASD | edge | 5 | ARIH2OS | 3 | 3.576e-06 | rs73077200 |
| ASD | edge | 6 | ARIH2 | 3 | 3.248e-06 | rs73077200 |
| ASD | edge | 7 | P4HTM | 3 | 2.545e-06 | rs73077200 |
| ASD | edge | 8 | WDR6 | 3 | 2.545e-06 | rs73077200 |
| ASD | edge | 9 | DALRD3 | 3 | 2.545e-06 | rs73077200 |
| ASD | edge | 10 | NDUFAF3 | 3 | 2.590e-06 | rs73077200 |
| ASD | edge | 11 | IMPDH2 | 3 | 1.178e-06 | rs73077200 |
| ASD | edge | 12 | QRICH1 | 3 | 1.066e-06 | rs73077200 |
| ASD | edge | 13 | QARS | 3 | 2.288e-06 | rs73077200 |
| ASD | edge | 14 | USP19 | 3 | 2.288e-06 | rs73077200 |
| ASD | edge | 15 | LAMB2 | 3 | 1.907e-06 | rs73077200 |
| ASD | edge | 16 | CCDC71 | 3 | 1.815e-06 | rs73077200 |
| ASD | edge | 17 | KLHDC8B | 3 | 1.815e-06 | rs73077200 |
| ASD | edge | 18 | C3orf84 | 3 | 1.815e-06 | rs73077200 |
| ASD | edge | 19 | CCDC36 | 3 | 1.730e-06 | rs73077200 |
| ASD | edge | 20 | RP11-3B7.1 | 3 | 2.966e-06 | rs73077200 |
| ASD | edge | 21 | C3orf62 | 3 | 2.913e-06 | rs73077200 |
| ASD | edge | 22 | USP4 | 3 | 2.913e-06 | rs73077200 |
| ASD | edge | 23 | RHOA | 3 | 4.940e-07 | rs73077200 |
| ASD | edge | 24 | AMT | 3 | 2.164e-06 | rs73077200 |
| ASD | edge | 25 | NICN1 | 3 | 2.164e-06 | rs73077200 |
| ASD | edge | 26 | DAG1 | 3 | 4.838e-07 | rs73077200 |
| ASD | edge | 27 | BSN | 3 | 1.755e-06 | rs73077200 |
| ASD | edge | 28 | MST1 | 3 | 6.134e-07 | rs73077200 |
| ASD | edge | 29 | RNF123 | 3 | 6.134e-07 | rs73077200 |
| ASD | edge | 30 | AMIGO3 | 3 | 2.019e-06 | rs73077200 |
| ASD | edge | 31 | GMPPB | 3 | 2.019e-06 | rs73077200 |
| ASD | edge | 32 | IP6K1 | 3 | 4.498e-07 | rs73077200 |
| ASD | edge | 33 | CDHR4 | 3 | 4.498e-07 | rs73077200 |
| ASD | edge | 34 | FAM212A | 3 |  | rs73077200 |
| ASD | edge | 35 | UBA7 | 3 | 8.581e-07 | rs73077200 |
| ASD | edge | 36 | TRAIP | 3 | 8.581e-07 | rs73077200 |
| ASD | edge | 37 | CAMKV | 3 | 1.480e-06 | rs73077200 |
| ASD | edge | 38 | RBM6 | 3 | 4.325e-06 | rs73077200 |
| ASD | edge | 39 | RBM5 | 3 |  | rs73077200 |
| ASD | edge | 40 | SEMA3F | 3 | 1.116e-06 | rs73077200 |
| ASD | edge | 41 | GNAT1 | 3 | 2.812e-06 | rs73077200 |
| ASD | edge | 42 | XKR6 | 8 | 9.265e-09 | rs11777007 |
| ASD | edge | 43 | PLEKHM1 | 17 | 2.088e-08 | rs71375338 |
| ASD | edge | 44 | CRHR1 | 17 | 1.166e-08 | rs71375338 |
| ASD | edge | 45 | SPPL2C | 17 | 9.595e-09 | rs71375338 |
| ASD | edge | 46 | MAPT | 17 | 9.713e-09 | rs71375338 |
| ASD | edge | 47 | STH | 17 | 1.470e-08 | rs71375338 |
| ASD | edge | 48 | KANSL1 | 17 | 7.877e-09 | rs71375338 |
| ASD | edge | 49 | ARL17B | 17 | 8.107e-09 | rs71375338 |
| ASD | edge | 50 | LRRC37A | 17 | 2.081e-08 | rs71375338 |
| ASD | edge | 51 | LRRC37A2 | 17 |  | rs71375338 |
| ASD | edge | 52 | ARL17A | 17 |  | rs71375338 |
| ASD | edge | 53 | NSF | 17 | 6.879e-08 | rs71375338 |
| ASD | node | 1 | PAPPA | 9 | 4.358e-06 | rs888398 |
| ASD | node | 2 | PAPPA-AS1 | 9 | 4.358e-06 | rs888398 |
| ASD | node | 3 | ASTN2 | 9 | 4.696e-07 | rs888398 |
| ASD | node | 4 | TRIM32 | 9 | 8.440e-07 | rs888398 |
| ASD | node | 5 | ARHGAP27 | 17 | 5.581e-07 | rs8070942 |
| ASD | node | 6 | PLEKHM1 | 17 | 2.615e-08 | rs8070942 |
| ASD | node | 7 | CRHR1 | 17 | 1.417e-08 | rs8070942 |
| ASD | node | 8 | SPPL2C | 17 | 1.261e-08 | rs8070942 |
| ASD | node | 9 | MAPT | 17 | 1.169e-08 | rs8070942 |
| ASD | node | 10 | STH | 17 | 1.806e-08 | rs8070942 |
| ASD | node | 11 | KANSL1 | 17 | 9.396e-09 | rs8070942 |
| ASD | node | 12 | ARL17B | 17 | 9.681e-09 | rs8070942 |
| ASD | node | 13 | LRRC37A | 17 | 2.606e-08 | rs8070942 |
| ASD | node | 14 | LRRC37A2 | 17 |  | rs8070942 |
| ASD | node | 15 | ARL17A | 17 |  | rs8070942 |
| ASD | node | 16 | NSF | 17 | 5.364e-08 | rs8070942 |
| ASD | node | 17 | WNT3 | 17 | 4.643e-08 | rs8070942 |
| ASD | node | 18 | FHOD3 | 18 | 3.336e-07 | rs58092984 |
| BIP3 | edge | 1 | PPM1M | 3 | 3.548e-06 | rs2267921 |
| BIP3 | edge | 2 | WDR82 | 3 | 3.548e-06 | rs2267921 |
| BIP3 | edge | 3 | DNAH1 | 3 | 1.894e-06 | rs2267921 |
| BIP3 | edge | 4 | BAP1 | 3 | 1.894e-06 | rs2267921 |
| BIP3 | edge | 5 | SEMA3G | 3 |  | rs2267921 |
| BIP3 | edge | 6 | TNNC1 | 3 | 4.191e-07 | rs2267921 |
| BIP3 | edge | 7 | NISCH | 3 | 4.191e-07 | rs2267921 |
| BIP3 | edge | 8 | CADM2 | 3 | 6.995e-08 | rs1248860 |
| BIP3 | edge | 9 | MSRA | 8 | 3.431e-08 | rs13282106; rs6984231 |
| BIP3 | edge | 10 | XPNPEP1 | 10 | 4.050e-06 | rs17127128 |
| BIP3 | edge | 11 | RP11-451M19.3 | 10 | 3.797e-06 | rs17127128 |
| BIP3 | edge | 12 | ADD3 | 10 | 6.356e-07 | rs17127128 |
| BIP3 | edge | 13 | TBC1D21 | 15 | 2.726e-06 | rs2045931 |
| BIP3 | edge | 14 | MBP | 18 | 3.256e-07 | rs8086634 |
| BIP3 | edge | 15 | DAND5 | 19 |  | rs1548546 |
| BIP3 | edge | 16 | NFIX | 19 | 2.333e-08 | rs1548546 |
| BIP3 | node | 1 | ADAMTSL4 | 1 | 3.069e-07 | rs12116492 |
| BIP3 | node | 2 | ADAMTSL4-AS1 | 1 | 3.069e-07 | rs12116492 |
| BIP3 | node | 3 | MCL1 | 1 | 3.436e-07 | rs12116492 |
| BIP3 | node | 4 | ENSA | 1 | 5.950e-07 | rs12116492 |
| BIP3 | node | 5 | GOLPH3L | 1 | 7.419e-07 | rs12116492 |
| BIP3 | node | 6 | HORMAD1 | 1 | 1.560e-06 | rs12116492 |
| BIP3 | node | 7 | CTSS | 1 | 1.584e-06 | rs12116492 |
| BIP3 | node | 8 | GCKR | 2 | 1.177e-06 | rs867282 |
| BIP3 | node | 9 | AC109829.1 | 2 | 8.798e-07 | rs867282 |
| BIP3 | node | 10 | C2orf16 | 2 | 9.320e-07 | rs867282 |
| BIP3 | node | 11 | ZNF512 | 2 | 6.213e-07 | rs867282 |
| BIP3 | node | 12 | CCDC121 | 2 | 8.658e-07 | rs867282 |
| BIP3 | node | 13 | GPN1 | 2 | 5.914e-07 | rs867282 |
| BIP3 | node | 14 | SUPT7L | 2 | 5.914e-07 | rs867282 |
| BIP3 | node | 15 | SLC4A1AP | 2 | 5.914e-07 | rs867282 |
| BIP3 | node | 16 | AC074091.13 | 2 | 2.928e-07 | rs867282 |
| BIP3 | node | 17 | MRPL33 | 2 | 9.847e-07 | rs867282 |
| BIP3 | node | 18 | AC110084.1 | 2 | 9.847e-07 | rs867282 |
| BIP3 | node | 19 | RBKS | 2 | 9.847e-07 | rs867282 |
| BIP3 | node | 20 | ANKRD44 | 2 | 2.401e-06 | rs35157131 |
| BIP3 | node | 21 | SF3B1 | 2 | 4.672e-07 | rs35157131 |
| BIP3 | node | 22 | COQ10B | 2 | 5.243e-07 | rs35157131 |
| BIP3 | node | 23 | HSPD1 | 2 | 5.036e-07 | rs35157131 |
| BIP3 | node | 24 | HSPE1 | 2 | 5.036e-07 | rs35157131 |
| BIP3 | node | 25 | HSPE1-MOB4 | 2 | 5.036e-07 | rs35157131 |
| BIP3 | node | 26 | MOB4 | 2 | 5.036e-07 | rs35157131 |
| BIP3 | node | 27 | RFTN2 | 2 | 5.594e-07 | rs35157131 |
| BIP3 | node | 28 | AC011997.1 | 2 | 1.756e-06 | rs35157131 |
| BIP3 | node | 29 | MARS2 | 2 | 1.811e-06 | rs35157131 |
| BIP3 | node | 30 | BOLL | 2 | 1.776e-06 | rs35157131 |
| BIP3 | node | 31 | PLCL1 | 2 | 2.003e-06 | rs35157131 |
| BIP3 | node | 32 | SATB2 | 2 | 4.495e-07 | rs260755 |
| BIP3 | node | 33 | STAB1 | 3 | 1.332e-07 | rs1010553 |
| BIP3 | node | 34 | NT5DC2 | 3 | 1.876e-07 | rs1010553 |
| BIP3 | node | 35 | SMIM4 | 3 | 1.876e-07 | rs1010553 |
| BIP3 | node | 36 | PBRM1 | 3 | 2.376e-07 | rs1010553 |
| BIP3 | node | 37 | GNL3 | 3 | 3.120e-07 | rs1010553 |
| BIP3 | node | 38 | GLT8D1 | 3 | 3.120e-07 | rs1010553 |
| BIP3 | node | 39 | SPCS1 | 3 | 3.416e-07 | rs1010553 |
| BIP3 | node | 40 | NEK4 | 3 | 2.984e-07 | rs1010553 |
| BIP3 | node | 41 | ITIH1 | 3 | 5.254e-07 | rs1010553 |
| BIP3 | node | 42 | ITIH3 | 3 | 1.044e-06 | rs1010553 |
| BIP3 | node | 43 | ITIH4 | 3 | 1.044e-06 | rs1010553 |
| BIP3 | node | 44 | RP5-966M1.6 | 3 | 1.044e-06 | rs1010553 |
| BIP3 | node | 45 | RP11-127H5.1 | 8 | 2.945e-07 | rs12545916 |
| BIP3 | node | 46 | ZFPM2 | 8 | 2.945e-07 | rs12545916 |
| BIP3 | node | 47 | SFXN2 | 10 | 2.558e-06 | rs284858 |
| BIP3 | node | 48 | WBP1L | 10 | 3.414e-07 | rs284858 |
| BIP3 | node | 49 | CYP17A1 | 10 | 1.177e-06 | rs284858 |
| BIP3 | node | 50 | C10orf32 | 10 | 1.031e-06 | rs284858 |
| BIP3 | node | 51 | C10orf32-ASMT | 10 | 1.031e-06 | rs284858 |
| BIP3 | node | 52 | AS3MT | 10 | 1.031e-06 | rs284858 |
| BIP3 | node | 53 | SMG6 | 17 | 4.878e-07 | rs12941621 |
| BIP3 | node | 54 | SRR | 17 | 5.503e-07 | rs12941621 |
| BIP3 | node | 55 | TSR1 | 17 | 2.272e-06 | rs12941621 |
| BIP3 | node | 56 | DAND5 | 19 |  | rs1548546 |
| BIP3 | node | 57 | NFIX | 19 | 1.412e-08 | rs1548546 |
| MDD | edge | 1 | HIST1H2BL | 6 | 2.552e-07 | rs6940116 |
| MDD | edge | 2 | HIST1H2AI | 6 | 2.552e-07 | rs6940116 |
| MDD | edge | 3 | HIST1H3H | 6 | 2.552e-07 | rs6940116 |
| MDD | edge | 4 | HIST1H2AJ | 6 | 4.904e-07 | rs6940116 |
| MDD | edge | 5 | HIST1H2BM | 6 | 4.904e-07 | rs6940116 |
| MDD | edge | 6 | HIST1H4J | 6 | 4.904e-07 | rs6940116 |
| MDD | edge | 7 | HIST1H4K | 6 | 5.570e-07 | rs6940116 |
| MDD | edge | 8 | HIST1H2AK | 6 | 5.570e-07 | rs6940116 |
| MDD | edge | 9 | HIST1H2BN | 6 | 5.570e-07 | rs6940116 |
| MDD | edge | 10 | HIST1H2AL | 6 | 2.043e-07 | rs6940116 |
| MDD | edge | 11 | HIST1H1B | 6 | 2.043e-07 | rs6940116 |
| MDD | edge | 12 | HIST1H3I | 6 | 2.043e-07 | rs6940116 |
| MDD | edge | 13 | HIST1H4L | 6 | 2.043e-07 | rs6940116 |
| SCZ | edge | 1 | FOXN2 | 2 | 1.310e-06 | rs3811519 |
| SCZ | edge | 2 | PPP1R21 | 2 | 3.568e-07 | rs3811519 |
| SCZ | edge | 3 | STON1 | 2 | 5.422e-06 | rs3811519 |
| SCZ | edge | 4 | STON1-GTF2A1L | 2 | 5.422e-06 | rs3811519 |
| SCZ | edge | 5 | VRK2 | 2 | 8.593e-09 | rs2717068 |
| SCZ | edge | 6 | FANCL | 2 | 1.395e-07 | rs2717068 |
| SCZ | edge | 7 | SLC4A10 | 2 | 2.499e-08 | rs2909457 |
| SCZ | edge | 8 | DPP4 | 2 | 2.499e-08 | rs2909457 |
| SCZ | edge | 9 | MLTK | 2 | 4.503e-07 | rs9636294 |
| SCZ | edge | 10 | CREB1 | 2 | 4.687e-07 | rs4675682 |
| SCZ | edge | 11 | METTL21A | 2 | 1.764e-06 | rs4675682 |
| SCZ | edge | 12 | TBC1D5 | 3 | 4.064e-08 | rs9855515 |
| SCZ | edge | 13 | C3orf49 | 3 | 3.405e-06 | rs9879045 |
| SCZ | edge | 14 | THOC7 | 3 | 3.405e-06 | rs9879045 |
| SCZ | edge | 15 | ATXN7 | 3 | 7.858e-07 | rs9879045 |
| SCZ | edge | 16 | PSMD6 | 3 | 4.208e-07 | rs9879045 |
| SCZ | edge | 17 | ZIC4 | 3 | 1.518e-07 | rs2279829 |
| SCZ | edge | 18 | ZIC1 | 3 | 1.518e-07 | rs2279829 |
| SCZ | edge | 19 | ELOVL7 | 5 | 7.272e-07 | rs10939881 |
| SCZ | edge | 20 | ERCC8 | 5 | 4.994e-07 | rs10939881 |
| SCZ | edge | 21 | NDUFAF2 | 5 | 2.188e-07 | rs10939881 |
| SCZ | edge | 22 | AC008498.1 | 5 | 2.971e-07 | rs10939881 |
| SCZ | edge | 23 | SMIM15 | 5 | 3.058e-07 | rs10939881 |
| SCZ | edge | 24 | HIST1H2BL | 6 | 5.471e-09 | rs2893931 |
| SCZ | edge | 25 | HIST1H2AI | 6 | 4.011e-09 | rs2893931 |
| SCZ | edge | 26 | HIST1H3H | 6 | 4.011e-09 | rs2893931 |
| SCZ | edge | 27 | HIST1H2AJ | 6 | 4.011e-09 | rs2893931 |
| SCZ | edge | 28 | HIST1H2BM | 6 | 4.011e-09 | rs2893931 |
| SCZ | edge | 29 | HIST1H4J | 6 | 4.011e-09 | rs2893931 |
| SCZ | edge | 30 | HIST1H4K | 6 | 3.475e-09 | rs2893931 |
| SCZ | edge | 31 | HIST1H2AK | 6 | 3.475e-09 | rs2893931 |
| SCZ | edge | 32 | HIST1H2BN | 6 | 3.475e-09 | rs2893931 |
| SCZ | edge | 33 | HIST1H3I | 6 | 2.662e-06 | rs2893931 |
| SCZ | edge | 34 | HIST1H4L | 6 | 2.662e-06 | rs2893931 |
| SCZ | edge | 35 | HIST1H3J | 6 | 2.662e-06 | rs2893931 |
| SCZ | edge | 36 | HIST1H2AM | 6 | 2.698e-06 | rs2893931 |
| SCZ | edge | 37 | HIST1H2BO | 6 | 2.698e-06 | rs2893931 |
| SCZ | edge | 38 | OR2B2 | 6 | 3.655e-06 | rs2893931 |
| SCZ | edge | 39 | TRIM27 | 6 | 1.097e-07 | rs880638 |
| SCZ | edge | 40 | C6orf100 | 6 | 4.587e-07 | rs880638 |
| SCZ | edge | 41 | ZNF311 | 6 | 5.918e-07 | rs880638 |
| SCZ | edge | 42 | MSRA | 8 | 7.098e-08 | rs7460436 |
| SCZ | edge | 43 | PRKG1 | 10 | 3.661e-07 | rs45506496 |
| SCZ | edge | 44 | DKK1 | 10 | 3.661e-07 | rs45506496 |
| SCZ | edge | 45 | MSRB3 | 12 | 3.488e-07 | rs10784456 |
| SCZ | edge | 46 | C12orf76 | 12 | 2.164e-06 | rs73194012 |
| SCZ | edge | 47 | IFT81 | 12 | 6.690e-07 | rs73194012 |
| SCZ | edge | 48 | ATP2A2 | 12 | 8.493e-07 | rs73194012 |
| SCZ | edge | 49 | ANAPC7 | 12 | 9.017e-07 | rs73194012 |
| SCZ | edge | 50 | ARPC3 | 12 | 7.307e-07 | rs73194012 |
| SCZ | edge | 51 | GPN3 | 12 | 7.080e-07 | rs73194012 |
| SCZ | edge | 52 | FAM216A | 12 | 6.640e-07 | rs73194012 |
| SCZ | edge | 53 | VPS29 | 12 | 6.497e-07 | rs73194012 |
| SCZ | edge | 54 | RAD9B | 12 | 6.445e-07 | rs73194012 |
| SCZ | edge | 55 | PPTC7 | 12 | 4.511e-07 | rs73194012 |
| SCZ | edge | 56 | TCTN1 | 12 | 3.130e-07 | rs73194012 |
| SCZ | edge | 57 | HVCN1 | 12 | 3.064e-07 | rs73194012 |
| SCZ | edge | 58 | PPP1CC | 12 | 5.541e-07 | rs73194012 |
| SCZ | edge | 59 | CCDC63 | 12 | 4.850e-06 | rs73194012 |
| SCZ | edge | 60 | DAAM1 | 14 | 1.457e-07 | rs77168169 |
| SCZ | edge | 61 | BCL11B | 14 | 4.433e-07 | rs2614464 |
| SCZ | edge | 62 | SMG6 | 17 | 9.735e-08 | rs11655813 |
| SCZ | edge | 63 | SRR | 17 | 9.108e-07 | rs11655813 |
| SCZ | edge | 64 | TSR1 | 17 | 4.483e-06 | rs11655813 |
| SCZ | edge | 65 | PLCD3 | 17 | 2.352e-07 | rs60289499 |
| SCZ | edge | 66 | ACBD4 | 17 | 2.352e-07 | rs60289499 |
| SCZ | edge | 67 | HEXIM1 | 17 | 2.352e-07 | rs60289499 |
| SCZ | edge | 68 | HEXIM2 | 17 | 4.399e-07 | rs60289499 |
| SCZ | edge | 69 | ARHGAP27 | 17 | 3.196e-06 | rs2532392 |
| SCZ | edge | 70 | PLEKHM1 | 17 | 6.841e-07 | rs2532392 |
| SCZ | edge | 71 | CRHR1 | 17 | 2.220e-07 | rs2532392 |
| SCZ | edge | 72 | SPPL2C | 17 | 2.236e-07 | rs2532392 |
| SCZ | edge | 73 | MAPT | 17 | 1.129e-07 | rs2532392 |
| SCZ | edge | 74 | STH | 17 | 1.806e-07 | rs2532392 |
| SCZ | edge | 75 | KANSL1 | 17 | 9.008e-08 | rs2532392 |
| SCZ | edge | 76 | ARL17B | 17 | 1.166e-07 | rs2532392 |
| SCZ | edge | 77 | LRRC37A | 17 |  | rs2532392 |
| SCZ | edge | 78 | LRRC37A2 | 17 |  | rs2532392 |
| SCZ | edge | 79 | ARL17A | 17 |  | rs2532392 |
| SCZ | edge | 80 | NSF | 17 | 2.444e-07 | rs2532392 |
| SCZ | edge | 81 | WNT3 | 17 | 1.688e-07 | rs2532392 |
| SCZ | edge | 82 | ATP5G1 | 17 | 1.713e-06 | rs11079848 |
| SCZ | edge | 83 | UBE2Z | 17 | 2.130e-06 | rs11079848 |
| SCZ | edge | 84 | SNF8 | 17 | 2.130e-06 | rs11079848 |
| SCZ | edge | 85 | GIP | 17 | 3.695e-07 | rs11079848 |
| SCZ | edge | 86 | DAND5 | 19 |  | rs8103241 |
| SCZ | edge | 87 | NFIX | 19 | 2.969e-07 | rs8103241 |
| SCZ | node | 1 | DPYD | 1 | 3.019e-06 | rs61789073 |
| SCZ | node | 2 | AKT3 | 1 | 2.167e-07 | rs12048930 |
| SCZ | node | 3 | GCKR | 2 | 1.322e-06 | rs12994085 |
| SCZ | node | 4 | AC109829.1 | 2 | 8.502e-07 | rs12994085 |
| SCZ | node | 5 | C2orf16 | 2 | 8.941e-07 | rs12994085 |
| SCZ | node | 6 | ZNF512 | 2 | 4.112e-07 | rs12994085 |
| SCZ | node | 7 | CCDC121 | 2 | 8.519e-07 | rs12994085 |
| SCZ | node | 8 | GPN1 | 2 | 3.871e-07 | rs12994085 |
| SCZ | node | 9 | SUPT7L | 2 | 3.871e-07 | rs12994085 |
| SCZ | node | 10 | SLC4A1AP | 2 | 3.871e-07 | rs12994085 |
| SCZ | node | 11 | AC074091.13 | 2 | 2.262e-07 | rs12994085 |
| SCZ | node | 12 | MRPL33 | 2 | 7.646e-07 | rs12994085 |
| SCZ | node | 13 | AC110084.1 | 2 | 7.719e-07 | rs12994085 |
| SCZ | node | 14 | RBKS | 2 | 7.646e-07 | rs12994085 |
| SCZ | node | 15 | VRK2 | 2 | 3.582e-07 | rs6722461 |
| SCZ | node | 16 | ANKRD44 | 2 | 3.774e-07 | rs35157131 |
| SCZ | node | 17 | SF3B1 | 2 | 2.911e-07 | rs35157131 |
| SCZ | node | 18 | COQ10B | 2 | 3.343e-07 | rs35157131 |
| SCZ | node | 19 | HSPD1 | 2 | 3.185e-07 | rs35157131 |
| SCZ | node | 20 | HSPE1 | 2 | 3.185e-07 | rs35157131 |
| SCZ | node | 21 | HSPE1-MOB4 | 2 | 3.185e-07 | rs35157131 |
| SCZ | node | 22 | MOB4 | 2 | 3.185e-07 | rs35157131 |
| SCZ | node | 23 | RFTN2 | 2 | 3.611e-07 | rs35157131 |
| SCZ | node | 24 | AC011997.1 | 2 | 1.446e-06 | rs35157131 |
| SCZ | node | 25 | MARS2 | 2 | 1.477e-06 | rs35157131 |
| SCZ | node | 26 | BOLL | 2 | 1.446e-06 | rs35157131 |
| SCZ | node | 27 | PLCL1 | 2 | 1.746e-06 | rs35157131 |
| SCZ | node | 28 | SATB2 | 2 | 2.762e-08 | rs17195804 |
| SCZ | node | 29 | GIGYF2 | 2 | 4.398e-07 | rs6704768 |
| SCZ | node | 30 | KCNJ13 | 2 | 1.463e-06 | rs6704768 |
| SCZ | node | 31 | C2orf82 | 2 | 1.730e-06 | rs6704768 |
| SCZ | node | 32 | NGEF | 2 | 3.592e-06 | rs6704768 |
| SCZ | node | 33 | STAB1 | 3 | 7.352e-08 | rs1010553 |
| SCZ | node | 34 | NT5DC2 | 3 | 1.049e-07 | rs1010553 |
| SCZ | node | 35 | SMIM4 | 3 | 1.049e-07 | rs1010553 |
| SCZ | node | 36 | PBRM1 | 3 | 1.354e-07 | rs1010553 |
| SCZ | node | 37 | GNL3 | 3 | 1.825e-07 | rs1010553 |
| SCZ | node | 38 | GLT8D1 | 3 | 1.825e-07 | rs1010553 |
| SCZ | node | 39 | SPCS1 | 3 | 2.025e-07 | rs1010553 |
| SCZ | node | 40 | NEK4 | 3 | 1.738e-07 | rs1010553 |
| SCZ | node | 41 | ITIH1 | 3 | 3.352e-07 | rs1010553 |
| SCZ | node | 42 | ITIH3 | 3 | 7.900e-07 | rs1010553 |
| SCZ | node | 43 | ITIH4 | 3 | 7.900e-07 | rs1010553 |
| SCZ | node | 44 | RP5-966M1.6 | 3 | 7.900e-07 | rs1010553 |
| SCZ | node | 45 | ZIC4 | 3 | 1.743e-07 | rs2279829 |
| SCZ | node | 46 | ZIC1 | 3 | 1.743e-07 | rs2279829 |
| SCZ | node | 47 | PPM1L | 3 | 3.934e-07 | rs9860738 |
| SCZ | node | 48 | B3GALNT1 | 3 | 2.655e-07 | rs9860738 |
| SCZ | node | 49 | NMD3 | 3 | 2.526e-07 | rs9860738 |
| SCZ | node | 50 | SPTSSB | 3 | 2.613e-06 | rs9860738 |
| SCZ | node | 51 | RNF175 | 4 | 1.038e-07 | rs10031057 |
| SCZ | node | 52 | SFRP2 | 4 | 9.426e-08 | rs10031057 |
| SCZ | node | 53 | KMT2E | 7 | 9.176e-07 | rs2470943 |
| SCZ | node | 54 | SRPK2 | 7 | 9.176e-07 | rs2470943 |
| SCZ | node | 55 | SGK223 | 8 | 1.752e-07 | rs2945899 |
| SCZ | node | 56 | STAR | 8 |  | rs1906672 |
| SCZ | node | 57 | LSM1 | 8 | 3.256e-07 | rs1906672 |
| SCZ | node | 58 | BAG4 | 8 | 3.716e-07 | rs1906672 |
| SCZ | node | 59 | DDHD2 | 8 | 1.497e-07 | rs1906672 |
| SCZ | node | 60 | PPAPDC1B | 8 | 1.497e-07 | rs1906672 |
| SCZ | node | 61 | WHSC1L1 | 8 | 1.497e-07 | rs1906672 |
| SCZ | node | 62 | LETM2 | 8 | 2.693e-07 | rs1906672 |
| SCZ | node | 63 | FGFR1 | 8 | 4.225e-07 | rs1906672 |
| SCZ | node | 64 | ZMAT4 | 8 | 2.390e-07 | rs62640287 |
| SCZ | node | 65 | MMP16 | 8 | 2.042e-07 | rs28630437 |
| SCZ | node | 66 | PIP5K1B | 9 | 3.494e-07 | rs4745405 |
| SCZ | node | 67 | SFXN2 | 10 | 5.723e-07 | rs284858 |
| SCZ | node | 68 | WBP1L | 10 | 1.757e-07 | rs284858 |
| SCZ | node | 69 | CYP17A1 | 10 | 9.197e-07 | rs284858 |
| SCZ | node | 70 | C10orf32 | 10 | 7.778e-07 | rs284858 |
| SCZ | node | 71 | C10orf32-ASMT | 10 | 7.778e-07 | rs284858 |
| SCZ | node | 72 | AS3MT | 10 | 7.778e-07 | rs284858 |
| SCZ | node | 73 | SORCS3 | 10 | 3.138e-07 | rs1027190 |
| SCZ | node | 74 | NTM | 11 | 3.054e-07 | rs7125824 |
| SCZ | node | 75 | TSPAN8 | 12 | 3.888e-07 | rs1463768 |
| SCZ | node | 76 | ABCB9 | 12 | 4.641e-08 | rs1626703 |
| SCZ | node | 77 | OGFOD2 | 12 | 4.641e-08 | rs1626703 |
| SCZ | node | 78 | ARL6IP4 | 12 | 4.641e-08 | rs1626703 |
| SCZ | node | 79 | PITPNM2 | 12 | 4.641e-08 | rs1626703 |
| SCZ | node | 80 | MPHOSPH9 | 12 | 4.286e-08 | rs1626703 |
| SCZ | node | 81 | C12orf65 | 12 | 4.286e-08 | rs1626703 |
| SCZ | node | 82 | CDK2AP1 | 12 | 9.905e-08 | rs1626703 |
| SCZ | node | 83 | SBNO1 | 12 | 7.436e-08 | rs1626703 |
| SCZ | node | 84 | SETD8 | 12 | 7.352e-08 | rs1626703 |
| SCZ | node | 85 | RILPL2 | 12 | 1.464e-07 | rs1626703 |
| SCZ | node | 86 | RBFOX1 | 16 | 1.481e-07 | rs810609 |
| SCZ | node | 87 | SMG6 | 17 | 7.895e-09 | rs9896535 |
| SCZ | node | 88 | SRR | 17 | 4.943e-08 | rs9896535 |
| SCZ | node | 89 | TSR1 | 17 | 1.363e-06 | rs9896535 |
| SCZ | node | 90 | EPN2 | 17 | 1.539e-09 | rs1467028 |
| SCZ | node | 91 | B9D1 | 17 | 2.214e-09 | rs1467028 |
| SCZ | node | 92 | MAPK7 | 17 | 1.529e-08 | rs1467028 |
| SCZ | node | 93 | MFAP4 | 17 | 1.529e-08 | rs1467028 |
| SCZ | node | 94 | ARHGAP27 | 17 | 3.399e-06 | rs2532392 |
| SCZ | node | 95 | PLEKHM1 | 17 | 6.479e-07 | rs2532392 |
| SCZ | node | 96 | CRHR1 | 17 | 1.613e-07 | rs2532392 |
| SCZ | node | 97 | SPPL2C | 17 | 1.629e-07 | rs2532392 |
| SCZ | node | 98 | MAPT | 17 | 6.873e-08 | rs2532392 |
| SCZ | node | 99 | STH | 17 | 1.241e-07 | rs2532392 |
| SCZ | node | 100 | KANSL1 | 17 | 5.208e-08 | rs2532392 |
| SCZ | node | 101 | ARL17B | 17 | 7.149e-08 | rs2532392 |
| SCZ | node | 102 | LRRC37A | 17 |  | rs2532392 |
| SCZ | node | 103 | LRRC37A2 | 17 |  | rs2532392 |
| SCZ | node | 104 | ARL17A | 17 |  | rs2532392 |
| SCZ | node | 105 | NSF | 17 | 1.825e-07 | rs2532392 |
| SCZ | node | 106 | WNT3 | 17 | 1.139e-07 | rs2532392 |
| SCZ | node | 107 | ZBTB14 | 18 | 3.636e-07 | rs1940942 |
| SCZ | node | 108 | DAND5 | 19 |  | rs8103241 |
| SCZ | node | 109 | NFIX | 19 | 2.338e-07 | rs8103241 |
| SCZ | node | 110 | SLC32A1 | 20 | 4.080e-07 | rs2263747 |
| SCZ | node | 111 | ACTR5 | 20 | 4.111e-07 | rs2263747 |
| SCZ | node | 112 | TOP1 | 20 | 2.561e-06 | rs17265513 |
| SCZ | node | 113 | PLCG1 | 20 | 2.956e-06 | rs17265513 |
| SCZ | node | 114 | ZHX3 | 20 | 3.337e-07 | rs17265513 |
| SCZ | node | 115 | LPIN3 | 20 | 7.542e-06 | rs17265513 |
| SCZ | node | 116 | GATSL3 | 22 | 1.196e-06 | rs13056976 |
| SCZ | node | 117 | RP1-130H16.18 | 22 | 1.007e-06 | rs13056976 |
| SCZ | node | 118 | TBC1D10A | 22 | 8.917e-07 | rs13056976 |
| SCZ | node | 119 | SF3A1 | 22 | 4.317e-07 | rs13056976 |
| SCZ | node | 120 | CCDC157 | 22 | 3.978e-07 | rs13056976 |
| SCZ | node | 121 | RNF215 | 22 | 5.369e-07 | rs13056976 |
| SCZ | node | 122 | SEC14L2 | 22 | 1.715e-06 | rs13056976 |
| SCZ | node | 123 | RP4-539M6.19 | 22 | 2.101e-06 | rs13056976 |
| SCZ | node | 124 | KIAA1658 | 22 | 2.101e-06 | rs13056976 |
| SCZ | node | 125 | MTFP1 | 22 | 2.101e-06 | rs13056976 |

**Suppl. Table 7. Mapped processes from SynGO.** Out of 180 genes identified through conjunctional FDR (Suppl. Table 5), 23 were linked to synaptic functioning by *SynGO*. The synaptic features are categorized in cellular components and biological processes. Gene symbols can appear multiple times in the table.

| **Gene symbol** | **Domain** | **Synaptic feature** | **Number of instances** |
| --- | --- | --- | --- |
| SYNGAP1 | Cellular component | Postsynaptic density, intracellular component (go:0099092) | 5 |
| DLG1 | Cellular component | Postsynaptic density, intracellular component (go:0099092) | 5 |
| DNM2 | Cellular component | Postsynaptic density, intracellular component (go:0099092) | 5 |
| INA | Cellular component | Postsynaptic density, intracellular component (go:0099092) | 5 |
| EXOC4 | Cellular component | Postsynaptic density, intracellular component (go:0099092) | 5 |
| INA | Cellular component | Postsynapse (go:0098794) | 4 |
| PPP1CC | Cellular component | Postsynapse (go:0098794) | 4 |
| ARFGEF2 | Cellular component | Postsynapse (go:0098794) | 4 |
| BCL11A | Cellular component | Postsynapse (go:0098794) | 4 |
| PLCG1 | Biological process | Modulation of chemical synaptic transmission (go:0050804) | 3 |
| STAU1 | Biological process | Modulation of chemical synaptic transmission (go:0050804) | 3 |
| SYNGAP1 | Biological process | Modulation of chemical synaptic transmission (go:0050804) | 3 |
| PPP1CC | Cellular component | Presynapse (go:0098793) | 2 |
| ARFGEF2 | Cellular component | Presynapse (go:0098793) | 2 |
| SLC6A4 | Cellular component | Integral component of postsynaptic membrane (go:0099055) | 2 |
| HCN1 | Cellular component | Integral component of postsynaptic membrane (go:0099055) | 2 |
| SLC6A4 | Cellular component | Integral component of presynaptic membrane (go:0099056) | 2 |
| NRXN1 | Cellular component | Integral component of presynaptic membrane (go:0099056) | 2 |
| NRXN1 | Cellular component | Integral component of presynaptic active zone membrane (go:0099059) | 2 |
| HCN1 | Cellular component | Integral component of presynaptic active zone membrane (go:0099059) | 2 |
| KIAA1109 | Biological process | Regulation of synaptic vesicle endocytosis (go:1900242) | 2 |
| VPS29 | Biological process | Regulation of synaptic vesicle endocytosis (go:1900242) | 2 |
| NRXN1 | Biological process | Synapse assembly (go:0007416) | 1 |
| EXOC4 | Cellular component | Synaptic vesicle (go:0008021) | 1 |
| SLC32A1 | Cellular component | Integral component of synaptic vesicle membrane (go:0030285) | 1 |
| YWHAE | Cellular component | Synapse (go:0045202) | 1 |
| DNM2 | Biological process | Synaptic vesicle endocytosis (go:0048488) | 1 |
| HCN1 | Biological process | Regulation of postsynaptic membrane potential (go:0060078) | 1 |
| NRXN1 | Biological process | Regulation of synaptic vesicle cycle (go:0098693) | 1 |
| CRK | Biological process | Postsynaptic specialization assembly (go:0098698) | 1 |
| SLC32A1 | Biological process | Synaptic vesicle neurotransmitter loading (go:0098700) | 1 |
| SLC6A4 | Biological process | Neurotransmitter reuptake (go:0098810) | 1 |
| SYNGAP1 | Biological process | Maintenance of postsynaptic specialization structure (go:0098880) | 1 |
| DLG1 | Biological process | Structural constituent of postsynaptic density (go:0098919) | 1 |
| STAU1 | Biological process | Dendritic transport of messenger ribonucleoprotein complex (go:0098963) | 1 |
| BDNF | Cellular component | Neuronal dense core vesicle (go:0098992) | 1 |
| STAU1 | Biological process | Modification of postsynaptic structure (go:0099010) | 1 |
| NRXN1 | Biological process | Presynapse assembly (go:0099054) | 1 |
| YWHAE | Biological process | Regulation of postsynaptic membrane neurotransmitter receptor levels (go:0099072) | 1 |
| TERF2 | Biological process | Axonal transport of messenger ribonucleoprotein complex (go:0099088) | 1 |
| NRXN1 | Biological process | Regulation of postsynaptic specialization assembly (go:0099150) | 1 |
| NRXN1 | Biological process | Regulation of postsynaptic density assembly (go:0099151) | 1 |
| INA | Cellular component | Postsynaptic intermediate filament cytoskeleton (go:0099160) | 1 |
| INA | Biological process | Postsynaptic modulation of chemical synaptic transmission (go:0099170) | 1 |
| BDNF | Biological process | Trans-synaptic signaling by bdnf, modulating synaptic transmission (go:0099183) | 1 |
| INA | Biological process | Structural constituent of postsynaptic intermediate filament cytoskeleton (go:0099184) | 1 |
| NSF | Biological process | Synaptic vesicle cycle (go:0099504) | 1 |
| NSF | Cellular component | Presynaptic cytosol (go:0099523) | 1 |
| DNM2 | Biological process | Presynaptic dense core vesicle exocytosis (go:0099525) | 1 |
| PLXNA4 | Biological process | Maintenance of synapse structure (go:0099558) | 1 |
| NRXN1 | Biological process | Synapse adhesion between pre- and post-synapse (go:0099560) | 1 |
| DLG1 | Biological process | Neurotransmitter receptor localization to postsynaptic specialization membrane (go:0099645) | 1 |
| HCN1 | Biological process | Intracellular camp-activated cation channel activity involved in regulation of presynaptic membrane potential (go:0140232) | 1 |
| CUL3 | Biological process | Regulation protein catabolic process at postsynapse (go:0140252) | 1 |
| NRXN1 | Biological process | Regulation of trans-synaptic signaling by endocannabinoid, modulating synaptic transmission (go:0150036) | 1 |
| NRXN1 | Biological process | Regulation of presynapse assembly (go:1905606) | 1 |

**Suppl. Table 8. Mapped processes from the reactome pathway database**. This table shows the biological processes associated with the genes from the conjunctional FDR analyses as detected by the reactome pathway database. Not all genes were mapped- Some genes are associated with multiple processes. Only pathways with p-value < 0.05 are shown.

| **Pathway** | **Number of entities found** | **Entities p-value** | **Genes involved** |
| --- | --- | --- | --- |
| M Phase | 11 | 0.027917412 | YWHAE, PPP1CC, ANAPC7, VPS4A, PSMD3, CENPL, VRK2, HIST1H2BD, HIST1H2AC, LPIN3, SDCCAG8 |
| TCF dependent signaling in response to WNT | 7 | 0.029301678 | TLE1, CUL3, PSMD3, CXXC4, HIST1H2BD, HIST1H2AC, WNT3 |
| Signaling by NTRKs | 6 | 0.026885145 | MAPK7, BDNF, FURIN, PLCG1, CRK, DNM2 |
| Chromosome Maintenance | 5 | 0.041340687 | CHTF8, CENPL, TERF2, HIST1H2BD, HIST1H2AC |
| DNA Damage/Telomere Stress Induced Senescence | 4 | 0.016637703 | HIST1H1E, TERF2, HIST1H2BD, HIST1H2AC |
| Diseases of programmed cell death | 4 | 0.035335336 | YWHAE, FOXO3, HIST1H2BD, HIST1H2AC |
| Senescence-Associated Secretory Phenotype (SASP) | 4 | 0.036556368 | MAPK7, ANAPC7, HIST1H2BD, HIST1H2AC |
| Anchoring of the basal body to the plasma membrane | 4 | 0.047194096 | YWHAE, TCTN1, B9D1, SDCCAG8 |
| Transcriptional Regulation by MECP2 | 4 | 0.048632439 | RBFOX1, BDNF |
| Packaging Of Telomere Ends | 3 | 0.011559968 | TERF2, HIST1H2BD, HIST1H2AC |
| TGF-beta receptor signaling activates SMADs | 3 | 0.013455651 | PPP1CC, FURIN |
| Recognition and association of DNA glycosylase with site containing an affected purine | 3 | 0.016612275 | TERF2, HIST1H2BD, HIST1H2AC |
| Recognition and association of DNA glycosylase with site containing an affected pyrimidine | 3 | 0.020148738 | TERF2, HIST1H2BD, HIST1H2AC |
| PRC2 methylates histones and DNA | 3 | 0.022718660 | PHF1, HIST1H2BD, HIST1H2AC |
| Cleavage of the damaged purine | 3 | 0.024067393 | TERF2, HIST1H2BD, HIST1H2AC |
| Elastic fibre formation | 3 | 0.025458620 | FBN2, MFAP4, FURIN |
| Depurination | 3 | 0.025458620 | TERF2, HIST1H2BD, HIST1H2AC |
| Inhibition of DNA recombination at telomere | 3 | 0.028368332 | TERF2, HIST1H2BD, HIST1H2AC |
| FOXO-mediated transcription of oxidative stress, metabolic and neuronal genes | 3 | 0.029886649 | FOXO3, PLXNA4 |
| Cleavage of the damaged pyrimidine | 3 | 0.033049616 | TERF2, HIST1H2BD, HIST1H2AC |
| Depyrimidination | 3 | 0.033049616 | TERF2, HIST1H2BD, HIST1H2AC |
| Deposition of new CENPA-containing nucleosomes at the centromere | 3 | 0.038107496 | CENPL, HIST1H2BD, HIST1H2AC |
| Nucleosome assembly | 3 | 0.038107496 | CENPL, HIST1H2BD, HIST1H2AC |
| Synthesis of PC | 3 | 0.039876259 | SLC44A2, LPIN3 |
| Activated NTRK2 signals through PLCG1 | 2 | 0.003136362 | BDNF, PLCG1 |
| MECP2 regulates transcription factors | 2 | 0.008407695 | RBFOX1 |
| MECP2 regulates transcription of neuronal ligands | 2 | 0.013836400 | BDNF |
| RUNX1 and FOXP3 control the development of regulatory T lymphocytes (Tregs) | 2 | 0.022840564 | IL2 |
| CRMPs in Sema3A signaling | 2 | 0.025382412 | FES, PLXNA4 |
| Sema3A PAK dependent Axon repulsion | 2 | 0.028033563 | FES, PLXNA4 |
| Protein repair | 2 | 0.028033563 | MSRA, MSRB3 |
| SEMA3A-Plexin repulsion signaling by inhibiting Integrin adhesion | 2 | 0.030790634 | FES, PLXNA4 |
| Pre-NOTCH Processing in Golgi | 2 | 0.039664591 | ATP2A2, FURIN |
| Signaling by NODAL | 2 | 0.046051265 | FURIN, FOXO3 |
| Activation and oligomerization of BAK protein | 1 | 0.026823591 | BAK1 |
| Defective MMAB causes MMA, cblB type | 1 | 0.039965726 | MMAB |
